# The genetic architecture of pain intensity in a sample of 598,339 U.S. veterans

**DOI:** 10.1101/2023.03.09.23286958

**Authors:** Sylvanus Toikumo, Rachel Vickers-Smith, Zeal Jinwala, Heng Xu, Divya Saini, Emily Hartwell, Mirko P. Venegas, Kyle A. Sullivan, Ke Xu, Daniel A. Jacobson, Joel Gelernter, Christopher T. Rentsch, Million Veteran Program, Eli Stahl, Martin Cheatle, Hang Zhou, Stephen G. Waxman, Amy C. Justice, Rachel L. Kember, Henry R. Kranzler

## Abstract

Chronic pain is a common problem, with more than one-fifth of adult Americans reporting pain daily or on most days. It adversely affects quality of life and imposes substantial personal and economic costs. Efforts to treat chronic pain using opioids played a central role in precipitating the opioid crisis. Despite an estimated heritability of 25-50%, the genetic architecture of chronic pain is not well characterized, in part because studies have largely been limited to samples of European ancestry. To help address this knowledge gap, we conducted a cross-ancestry meta-analysis of pain intensity in 598,339 participants in the Million Veteran Program, which identified 125 independent genetic loci, 82 of which are novel. Pain intensity was genetically correlated with other pain phenotypes, level of substance use and substance use disorders, other psychiatric traits, education level, and cognitive traits. Integration of the GWAS findings with functional genomics data shows enrichment for putatively causal genes (n = 142) and proteins (n = 14) expressed in brain tissues, specifically in GABAergic neurons. Drug repurposing analysis identified anticonvulsants, beta-blockers, and calcium-channel blockers, among other drug groups, as having potential analgesic effects. Our results provide insights into key molecular contributors to the experience of pain and highlight attractive drug targets.

## Introduction

Pain is an unpleasant sensory and emotional experience associated with, or resembling that associated with, actual or potential tissue damage^1^. Pain is often classified as either acute, which typically lasts less than 4 weeks, and chronic, lasting more than three months and potentially maladaptive^2^. An individual’s experience of pain is influenced by biological, psychological, and social factors^1,3^.

In a national survey, 50.2 million US adults (20.5%) reported experiencing pain on most days or every day^4^, making pain the most common reason for seeking medical treatment^5^ and resulting in total healthcare costs of 560 to 635 billion dollars in 2010^6^. Chronic pain is also associated with a poor quality of life^7^. In the late 1980’s many medical and pain organizations adopted policies to increase patients’ access to pain management, including opioids. These policies included efforts to ensure the adequate assessment of pain, which was designated as “the fifth vital sign”^2^. The resulting dramatic increase in prescriptions for opioid analgesics contributed to the opioid epidemic and a doubling of opioid-related deaths in the 1990s^8^’^9^.

Success rates for treating chronic pain with currently available medications are estimated to be as low as 10%^10^. Opioids are not efficacious in managing chronic non-cancer pain^11^ and their long-term use is associated with adverse effects such as addiction, sleep disturbance, opioid-induced hyperalgesia, endocrine changes, and cardiac and cognitive effects^12,13^. Other medications used to treat chronic noncancer pain, such as non-steroidal anti-inflammatory medications and antiepileptic drugs, are effective for only some types of pain and may be associated with significant adverse effects^14^. Because non-pharmacologic interventions are not accessible to most patients with pain, safe and efficacious medications are needed to address this highly prevalent condition. Thus, novel therapeutic targets for chronic pain are needed to facilitate the discovery or repurposing of safe, effective analgesics.

Notably, drug development efforts informed by genetics can double the rate of success^15-17^. Although the héritability (h^2^) of individual differences in the susceptibility to develop chronic pain is estimated in twin and family studies to be 25–50%^18,19^, the mechanisms that underlie it are poorly understood^20^. To date, genome-wide association studies (GWAS) of chronic pain in large samples, including the UK Biobank (UKBB) and 23andMe cohorts, have focused on specific bodily sites^21-24^ or aspects of an individual’s sensitivity to experiencing and reporting pain^25-28^. Although in samples of 150,000 to nearly 500,000 individuals GWAS have identified genome-wide significant (GWS) loci for headache^29^, osteoarthritis^30,31^, low back pain^23,24^, knee pain^21^, neuropathic pain^32^, and multisite chronic pain^25,26^, they have yielded few overlapping loci. This may be due to the different pain phenotypes employed, despite their having high genetic correlations among them^27,33^.

There are also significant genetic correlations between pain phenotypes and psychiatric, substance use, cognitive, anthropometric, and circadian traits^21,23-25,29,34^. This shared genetic predisposition suggests that a common genetic susceptibility underlies a broad range of diverse chronic pain conditions^34^ and common co-occurring conditions. For example, Mendelian randomization (MR) and latent causal variable analyses have shown positive causal effects of specific bodily site pain on depression^35,36^ and bi-directional casual associations between multisite chronic pain and major depressive disorder (MDD)^25,35^.

Despite a growing literature on pain GWAS, most studies have been conducted in predominantly European ancestry cohorts recruited from non-clinical biobanks. However, biobanks linked to electronic health records (EHRs) with large, well-characterized, multi-ancestry samples are now available for use in identifying genetic risk factors and therapeutic targets for chronic pain^37^. The Million Veteran Program (MVP)^38^, an observational cohort study and mega-biobank implemented in the U.S. Department of Veterans Affairs (VA) health care system, includes data on routine pain screening. Pain ratings in the MVP use an 11-point ordinal Numeric Rating Scale (NRS), which has been a standard practice in VA primary care for more than a decade^39^. The NRS has been shown to be a consistent, valid measure of reported pain^40-42^ and is particularly informative for a GWAS of pain, as over 50% of VA patients experience chronic pain^43^.

We conducted a cross-ancestry meta-analysis of the NRS in samples of African American (AA), European American (EA) and Hispanic American (HA) ancestries from the MVP (N = 598,339). Because of the frequency with which the NRS is administered to patients in the VA, for each individual we calculated the median annual score and then the median across years. Thus, although the NRS is a report of pain intensity experienced at a specific point in time, the median of medians provided a proxy for chronic pain. We also conducted a secondary analysis in a subsample of 566,959 individuals that excluded participants with a lifetime opioid use disorder (OUD) diagnosis to assess potential confounding by OUD.

## Methods

### Overview of analyses

We conducted ancestry-specific GWASs of pain scores using an 11-point ordinal NRS in a) all AAs, EAs, and HAs with pain ratings from the MVP and b) a subset of these participants that excluded those with a lifetime OUD diagnosis, each followed by a cross-ancestry meta-analysis. Details on phenotyping are provided below. Downstream analyses are based principally on the GWAS of pain scores in the full sample, complemented by the estimated heritability and genetic correlations (*r,s*) for the sample exclusive of participants with OUD. An overview of the analyses is provided in Supplementary Fig. 1.

### Million Veteran Program cohort

The MVP^38^ is an EHR-based cohort comprising >900,000 veterans recruited at 63 VA medical centers nationwide. All participants provided written informed consent, a blood sample for DNA extraction and genotyping, and approval to securely access their EHR for research purposes. The protocol and consent were approved by the Central Veterans Affairs Institutional Review Board (IRB) and all site-specific IRBs. All relevant guidelines for work with human participants were followed in the conduct of the study.

### Phenotype description

As early as 2000, the VA recommended using the NRS to routinely measure pain in clinical practice as a “fifth vital sign”^44^. Since that time, veterans have been asked to rate their pain severity in response to the question: “Are you in pain?” They then rated their current pain on a scale of 0-10 where “0 is no pain and 10 is the worst pain imaginable”. Participants had at least one inpatient or outpatient pain rating in the EHR. We included 598,339 individuals with 76,798,104 NRS scores (median number of scores = 109, IQR = 28 – 351) in the primary GWAS. To reduce the large number of pain observations, we calculated the median pain score by year for each participant and the median of the annual median pain scores. In a supplementary GWAS we excluded individuals with a documented ICD-9/10 diagnosis code for OUD in the EHR, yielding a total of 566,959 study participants. Demographic characteristics for the supplementary sample are presented in Supplementary Table 1.

### Genotyping and imputation

DNA samples were genotyped on the Affymetrix Axiom Biobank Array (MVP Release 4). For genotyped SNPs, standard quality control (QC) and subsequent imputation were implemented. Full details about SNP and sample QC by the MVP Genomics Working Group are published^45^. Briefly, DNA samples were removed for sex mismatch, having seven or more relatives in MVP (kinship > 0.08), excessive heterozygosity, or genotype call rate < 98.5%. Variants were removed if they were monomorphic, had a high degree of missingness (call rate < 0.8) or a Hardy–Weinberg equilibrium (HWE) threshold of *P*⍰<⍰1⍰×⍰10^−6^ both in the entire sample using a principal-component analysis (PCA)- adjusted method and within one of the three major ancestral groups (AA, EA and HA).

Genotype phasing and imputation were performed using SHAPEIT4 (v.4.1.3)^46^ and Minimac4 software^47^, respectively. Biallelic SNPs were imputed using the African Genome Resources reference panel by the Sanger Institute (comprising all samples from the 1000 Genomes Project phase 3, version 5 reference panel^48^, and 1,500 unrelated pan-African samples). Non-biallelic SNPs and indels were imputed in a secondary imputation step using the 1000 Genomes Project phase 3, version 5 reference panel^48^, with indels and complex variants from the second imputation merged into the African Genome Resources imputation.

We removed one individual from each pair of related individuals (kinship⍰>⍰0.08, *N* ⍰=⍰31,010) at random. The HARE method^49^ was used to classify subjects into major ancestral groups (AA = 112,968, EA = 436,683, HA = 48,688) and QC of imputed variants was performed within each ancestral group. SNPs with imputation quality (INFO) score ⍰<⍰0.7; minor allele frequency (MAF) in AAs⍰>⍰O.005, EAsEMO.001, and Has⍰> ⍰O. 01; a genotype call rate⍰ > ⍰O.95; or an HWE *P*⍰>⍰1⍰×⍰10^−6^were excluded.

### Association analyses and risk locus definition

Genome-wide association testing was based on a linear regression model using PLINK (v.2.O)^50^ and was adjusted for sex, age at enrollment, and the first 10 within-ancestry genetic principal components (PCs). Due to substantial differences in sample size across ancestral groups, meta-analyses were performed using a sample-size weighted method in METAL^51^. Variants with *P*⍰>⍰5⍰×⍰10^−8^ were considered genome-wide significant (GWS).

To identify risk loci and their lead variants, we performed LD clumping in FUMA^52^ at a range of 3,000 kb, r^2^ > 0.1, and the respective ancestry 1000 Genomes reference panel^48^. Following clumping, genomic risk loci within 1 Mb of one another were incorporated into the same locus. We used GCTA COJO^53^ to define independent variants by conditioning them on the most significant variant within the locus. After conditioning, significant variants (*P*⍰>⍰5⍰×⍰10^−8^) were considered independently associated. We performed a sign test to compare the direction of SNP effects across individual ancestral datasets. Independent lead variants in EAs were examined in AAs and HAs and a binomial test used to evaluate the null hypothesis that 50% of variants have the same effect direction across ancestries. For lead SNPs in EAs that were absent in AAs and HAs, we considered proxy GWS SNPs (*p* < 5×10^−8^) in high LD with the EA lead variant (*r*^2^ >0.8).

To prioritize credible sets of variants driving our GWAS results, we used FINEMAP^54^ to fine-map regions defined by LD clumps (r^2^ > 0.1). Because fine-mapping requires data from all markers in the region of interest^55^, we merged LD clumps that physically overlapped (within a 1-MB window of the lead variant) and excluded SNPs in the major histocompatibility complex (MHC) region due to its complexity. FINEMAP credible set reports the likelihood of causality using the marginal posterior probability (PP), which ranges from 0 to 1, with values closer to 1 being most likely causal.

### SNP-based heritability and functional enrichment

We used the linkage disequilibrium score (LDSC) regression^56^ method to estimate the SNP-based heritability (h^2^_SNP_) of pain intensity (in both the full and the supplementary samples) in AAs and EAs based on common SNPs in HapMap3^57^. Due to the small HA sample size, we could not calculate h^2^_SNP_ in this population. To ensure matching of population LD structure, pre-calculated LD scores for EAs were derived from the 1000 Genomes European reference population (version 3)^49^ using LDSC^56^. In-sample LD scores for AAs were calculated from MVP AA genotype data using cov-LDSC^58^.

We used S-LDSC to partition the SNP heritability for pain intensity among EAs and explored the enrichment of the partitioned heritability by functional genomic categories^59,60^ using three models: (a) a baseline-LD model that contains 75 overlapping annotations, including coding and regulatory regions of the genome and epigenomic features^59^ (b) a specific tissue model that examines 10 overlapping celltype groups derived from 220 cell-type-specific histone marks, including methylated histone H3 Lys4 (H3K4me1), trimethylated histone H3 Lys4 (H3K4me3), acetylated histones H3 Lys4 (H3K4ac) and H3K27ac^59^ and (c) a multi-tissue model based on gene expression and chromatin datasets generated by GTEx^61^ and the Roadmap Epigenomics Mapping Consortium^62^. For each model, we excluded multi-allelic and MHC region variants. Functional categories within each model were considered significantly enriched based on a Bonferroni-corrected *P* value.

### Gene-set functional characterization

We applied multi-marker analysis of genomic annotation (MAGMA) v.1.08^63^ in FUMA (v1.3.6a)^52^ to identify genes and gene sets associated with the findings from the pain intensity GWAS and metaanalysis. Using the default setting in MAGMA, we mapped GWS SNPs to 18,702 protein-coding genes according to their physical position in NCBI build 37. We also used chromatin interaction (Hi-C) coupled MAGMA (H-MAGMA)^64^ to assign non-coding (intergenic and intronic) SNPs to genes based on their chromatin interactions. H-MAGMA uses six Hi-C datasets derived from fetal brain, adult brain (N = 3), induced pluripotent stem cell (iPSC)-derived neurons and iPSC-derived astrocytes^65^. We applied a Bonferroni correction (MAGMA, *α* = 0.05/18,702; H-MAGMA, α = 0.05/293157/6) to identify genes significantly associated with pain intensity, correcting for all genes tested in each analysis (see Supplementary Tables 15 and 21 for full lists).

To determine the plausible tissue enrichment of mapped genes, we integrated our cross-ancestry and EA GWAS results with gene expression data from 54 tissues (GTE × v8) in FUMA^52^. Next, we used FUMA to curate gene sets and Gene Ontology terms (from the Molecular Signature Database v.7.0^66^). We corrected for gene size, density of variants, and LD pattern between genes in each tissue (Bonferroni-corrected *α* = 0.05/54).

Enrichment for cell-type specific (CTS) transcriptomic profiles was performed in FUMA^67^ using 13 human single-cell RNA-sequencing (sc-RNAseq) datasets derived from brain (see Supplementary Table 14 for a detailed list). FUMA estimates CTS transcriptomic enrichment from the sc-RNAseq in three ways: (1) per selected dataset, (2) within datasets using a conditionally independent analysis (based on stepwise conditional testing of *P* values for each cell type that passes Bonferroni correction within the same dataset), and (3) across datasets (testing for proportional significance across the results from step 2). Proportional significance (PS) reports the confidence level for observed cell type enrichment as low significance: < 0.5, jointly significant: 0.5 − 0.8; and independently significant: > 0.8. We considered CTS enrichments with conditional independent signals (*P* < 0.05) and PS > 0.5 to be driven by joint/independent genetic signals in our pain intensity GWAS results.

### Transcriptomic and proteomic regulation

To identify genes and proteins whose expression is associated with pain intensity, we integrated EA GWAS results with human brain transcriptomic (eQTL, N = 452; and sQTL, N = 452)^68,69^ and proteomic (N = 722)^62^ data. We also obtained pretrained models of gene expression from GTEx v.8 for five brain tissues significantly enriched in MAGMA analyses – cerebellum, cerebellar hemisphere, cortex, frontal cortex, and anterior cingulate cortex^61,71^. Human brain transcriptomic and proteomic data for dorsolateral prefrontal cortex were derived from the study by Wingo et al^70^. Transcriptome-wide association study (TWAS) and proteome-wide association study (PWAS) analyses were performed using the FUSION pipeline^71^ with Bonferroni correction (α = 0.05/N genes tested) to account for multiple testing.

We used the colocalization (coloc R package^72^ in FUSION^71^) as our primary method to identify SNPs that mediate association with pain intensity through effects on gene and protein expression and a posterior colocalization probability (PP) of 80% to denote a shared causal signal. To test the robustness of the colocalized signals, we also performed summary-based Mendelian randomization (SMR) analyses^73^. We applied the HEIDI test^73^ to filter out SMR signals (*P*HEIDI < 0.05) due to linkage disequilibrium between pain-associated variants and eQTLs/sQTLs. Human brain cis-eQTL and cis-sQTL summary data were obtained from Qi et al^74^ and GTEx^61^. For genomic regions containing multiple genes with significant SMR associations, we selected the top-associated cis-eQTL. We used Bonferroni correction to correct for multiple testing (α = 0.05/N genes tested).

To explore the enrichment of causal genes and proteins in the dorsal root ganglia (DRG), we accessed human and mouse RNA-seq data from 13 tissues (6 neural and 7 non-neural) from the DRG sensoryomics repository^75^. The data contain relative gene abundances in standardized transcripts per million mapped reads and have been normalized to allow comparison across genes. The proportions of gene expression in the CNS (neural proportion score) and DRG (DRG enrichment score) in the context of profiled tissues were calculated, as described in Ray et al^75^. Scores ranging from 0 to 1 were used to denote the strength of tissue enrichment.

### Drug repurposing

We examined the drug repurposing status of genes in EAs (N = 156) with high causal probability from fine mapping and transcriptomic and proteomic analyses, using the Druggable Genome database^76^. For completeness, we also included the significantly associated genes mapped to GWS variants and MAGMA results in AAs (N = 7) and HAs (N = 2). The Druggable Genome database contains 4,479 coding gene sets with the potential to be modulated by a drug-like small molecule based on their nucleotide sequence and structural similarity to targets of existing drugs^76^. This druggable genome was divided into three tiers. Tier 1 (N = 1,427) contains targets of licensed small molecules and biotherapeutic drugs (curated from the ChEMBL database^77^) and drugs in clinical development. Tier 2 (N = 682) includes targets with verified bioactive drug-like small molecule binding partners and > 50% identity with approved drug targets based on their nucleotide sequence. Tier 3 (N = 2,370) comprises targets or secreted proteins with more distant similarity with an approved drug and members of active protein complexes not included in Tiers 1 and 2. All causal genes and those reported in any of the three tiers of the Druggable Genome were also examined for interaction with prescription drug targets in clinical development using the Drug-Gene Interaction database (DGIdb)^78^, which compiles clinical trial information from the FDA, PharmGKB, Therapeutic Target Database, and DrugBank databases, among others. We categorized each prescription drug identified using the Anatomical Therapeutic Chemical classification system, retrieved from the Kyoto Encyclopedia of Genes and Genomics (https://www.genome.jp/kegg/drug/).

### Genetic correlation

We used LDSC^56^ to calculate the r_g_ of pain intensity with (a) 89 other published pain, substance use, medication use, psychiatric, and anthropometric traits from EA datasets selected using prior epidemiological evidence and (b) 12 psychiatric, substance use, and anthropometric traits based on available AA GWAS summary data (see Supplementary Tables 24 and 26 for detailed lists). In EAs, all traits were tested using pre-computed LD scores for HapMap3^57^, while in AAs, LD scores derived using cov-LDSC^58^ from MVP AA genotype data were used. In a hypothesis-neutral manner, we also calculated *r*_g_s of pain intensity with 1344 published and unpublished traits from the UKBB using the Complex Trait Virtual Lab (CTG-VL) (https://genoma.io/). CTG-VL is a free open-source platform that incorporates publicly available GWAS data that allow for the calculation of *r*_g_ for complex traits using LDSC^79^. Each set of *r*_g_ analyses was Bonferroni corrected to control for multiple comparisons (α = 0.05/number of traits tested).

We also estimated the cross-ancestry *r*_*g*_s for pain intensity between AAs, EAs and HAs using Popcorn^80^, a computational method that determines the correlation of causal-variant effect sizes at SNPs common across population groups using GWAS summary-level data and LD information. Ancestry-specific LD scores were derived from the 1000 Genomes reference population^48^.

### Polygenic risk score-based phenome-wide association studies

We calculated polygenic risk scores (PRS) for pain intensity and performed a PheWAS analysis in two samples - the Yale-Penn sample and the Penn Medicine Biobank (PMBB). The Yale-Penn sample^81^ was deeply phenotyped using the Semi-Structured Assessment for Drug Dependence and Alcoholism (SSADDA), a comprehensive psychiatric instrument that assesses physical, psychosocial, and psychiatric aspects of SUDs and comorbid psychiatric traits^82,83^. As described in detail previously^81^, genotyping was performed using the Illumina HumanOmni1-Quad microarray, the Illumina HumanCoreExome array, or the Illumina Multi-Ethnic Global array, followed by imputation using Minimac3^84^ and the 1000 Genomes Project phase3 reference panel^48^ implemented on the Michigan imputation server (https://imputationserver.sph.umich.edu). SNPs with imputation quality (INFO) score⍰<⍰0.7, MAF < 0.01, missingness > 0.01, or an allele frequency difference between batches > 0.04; and individuals with genotype call rate⍰<⍰30.95, or related individuals with pi-hat > 0.25 were excluded. PCs were used to determine genetic ancestry based on the 1000 Genomes Project phase3^48^. The resulting dataset included 4,922 AAs and 5,709 EAs.

The PMBB^85^ is linked to EHR phenotypes. PMBB samples were genotyped with the GSA genotyping array. Genotype phasing was done using EAGLE^84^ and imputation was performed using Minimac3^84^ on the TOPMed Imputation server^47^. Following QC (INFO< 0.3, missingness >0.95, MAF > 0.5, sample call rate > 0.9), PLINK 1.90 was used to identify and remove related individuals based on identity by descent (Pi-hat > 0.25). To estimate genetic ancestry, PCs were calculated using SNPs common to the PMBB and the 1000 Genomes Project phase3^48^ and the smartpea module of the Eigensoft package (https://github.com/DReichLab/EIG). Participants were assigned to an ancestral group based on the distance of 10 PCs from the 1000 Genomes reference populations. The resulting dataset included 10,383 AAs and 29,355 EAs.

PRSs for pain intensity were calculated in the Yale-Penn and the PMBB datasets using PRS-Continuous shrinkage software (PRS-CS)^86^, with the default setting used to estimate the shrinkage parameters and the random seed fixed to 1 for reproducibility. To identify associations between the pain intensity PRSs and phenotypes, we performed a PheWAS in each dataset by fitting logistic regression models for binary traits and linear regression models for continuous traits. Analyses were conducted using the PheWAS v0.12 R package^87^ with adjustment for sex, age at enrollment (in PMBB) or at interview (in Yale-Penn) and the first 10 PCs within each genetic ancestry. We Bonferroni corrected each ancestry-specific analysis (Yale-Penn EAs and AAs: *P* <8.10 × 10^−5^, PMBB EAs and AAs: *P <* 3.68 × 10^− 5^).

### Mendelian Randomization

We used two-sample Mendelian randomization^88^ to evaluate causal associations between 16 genetically correlated traits and pain intensity among EAs only because the two other population groups provided inadequate statistical power for the analysis. We inferred causality bidirectionally using three methods: weighted median, inverse-variance weighted (IVW) and MR-Egger, followed by a pleiotropy test using the MR Egger intercept. Instrumental variants were associated with the exposure at *P*⍰<⍰1⍰×⍰MIO^− 5^ and a clumping threshold of *r*^2^ = 0.01. Potential causal effects were those for which at least two MR tests were significant after multiple correction *(P* = 3.13EM10^− 3^, 0.05/16) and did not violate the assumption of horizontal pleiotropy (MR-Egger intercept *P*⍰>⍰0.05).

## Results

### Description of the sample

The study sample comprised 598,339 individuals (AA = 112,968, EA = 436,683, HA = 48,688), of whom 91.2% were male (Supplementary Table 1). The supplementary analyses from which individuals with a lifetime OUD diagnosis were excluded were reduced by 5% across population groups (AA = 104,050, EA = 415,740, HA = 46,169) (Supplementary Table 1). The median ages were 61.4 (s.d = 14.0) and 61.7 (s.d = 14.1) in the full and supplementary samples, respectively. About half of individuals in both the full sample (51.2%) and the supplementary sample (52.7%) reported a median NRS of 0, i.e., no pain. Mild (NRS 1-3), moderate (NRS 4-6) and severe pain (NRS 7-10) were reported by 24.4%, 19.2%, and 4.5%, respectively in the full sample, and 24.6%, 18.2%, and 4.0%, respectively in the supplementary sample.

### Identification of pain intensity risk loci

In our cross-ancestry meta-analysis of AA, EA, and HA samples, we identified 4,416 GWS variants represented by 158 LD-clumped index variants (r^2^ > 0.1) (Figure 1). Analyses conditioned on the lead SNP left 125 independent association signals (Supplementary Table 2), 42 of which have previously been reported as pain-related loci^23,25^ and 82 of which are novel (Supplementary Table 2). Eight independent variants are exonic, 84 reside within a gene transcript, and 33 are intergenic. Of the 8 exonic variants, 2 have likely damaging (PolyPhen > 0.5, CADD > 15) effects (S/.C39A8-rs13107325 and *WSCD2*-rs3764002) and 5 are potentially deleterious (CADD > 15; *ANAPC4*-rs34811474, *MIER*-rs2034244, *NUCB2*-rs757081, *AKAP10*-rs203462 and *APOF*-rs429358) (Supplementary Table 2). The GWAS in EAs yielded 103 LD clumps (r^2^ > 0.1) across 86 independent loci (Supplementary Figure 2, Supplementary Table 3). Of these, 15 were not GWS in the cross-ancestry meta-analysis (Supplementary Table 3). We also identified 2 GWS variants in 1 locus (nearest gene *PPARD;* chr 6) in AAs, and 15 GWS variants in 2 loci (nearest genes *RNU6-461P;* chr 3 and *RNU6-741P;* chr 15) in HAs (Supplementary Table 4).

**Figure 1.**
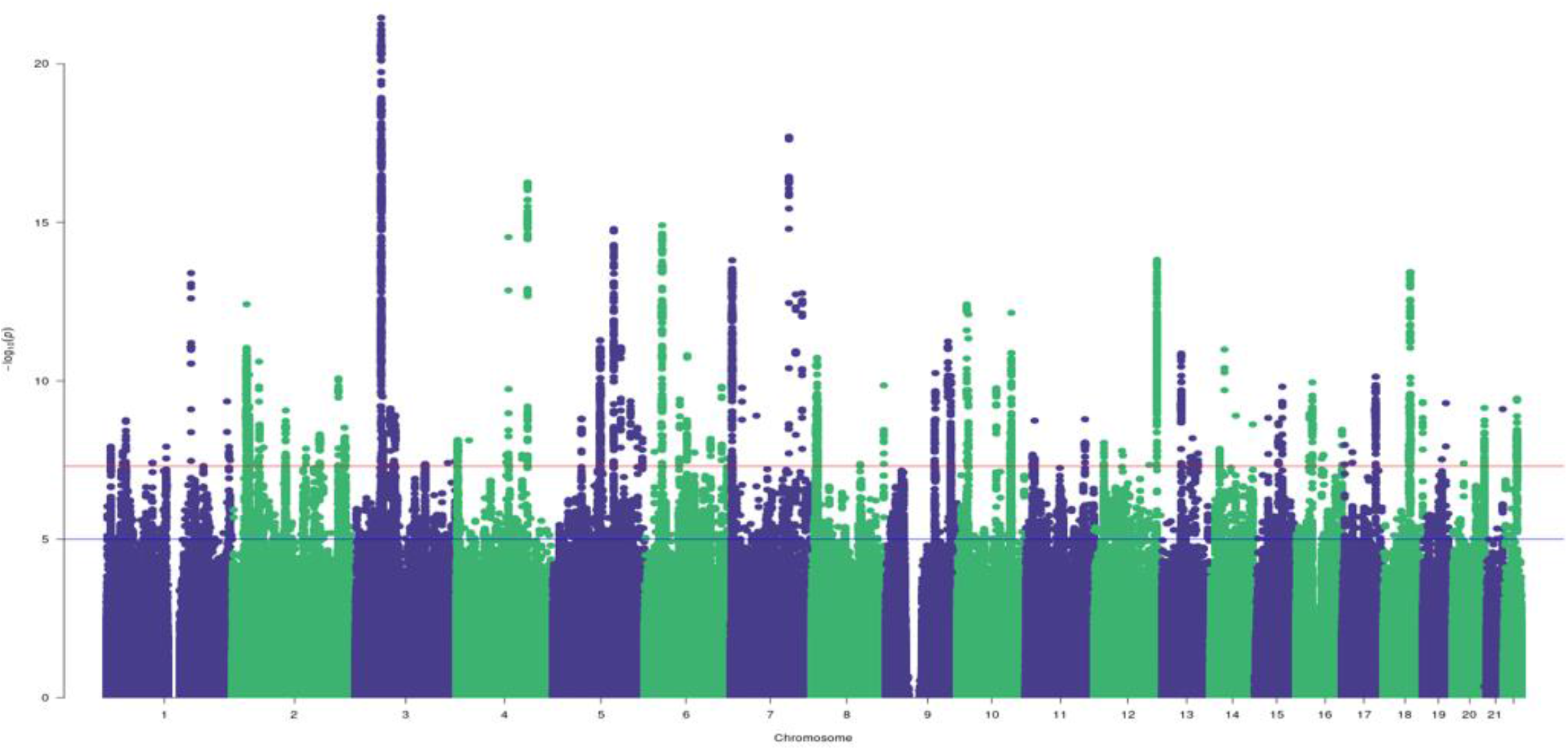
Manhattan plot for the pain intensity cross-ancestry GWAS meta-analysis. This identified 125 independent index variants. SNPs above the red line are GWS after correction for multiple testing (*P*⍰<⍰5⍰X⍰10^−8^)

We used a sign test to examine the 86 independent EA index variants in AAs and HAs, of which 57 and 74, respectively, were directly analyzed or had proxy SNPs in these populations (Supplementary Table 5). Most variants had the same direction of effect in both populations (*N*_SNPs_ AAs = 41, HAs = 61; sign test AAs *P* =⍰.0013, HAs *P* =⍰1.39⍰×⍰1O^−8^). Only 15 variants (*N*_SNPS_ AAS = 2, HAs = 13) were nominally associated (*P* <⍰0.05) and none survived multiple test correction (Supplementary Table 5). The cross-ancestry genetic-effect correlation (ρ_pe_) was 0.71 (SE = 0.13, *P =* 2.12 × 10^−2^) between EAs and AAs and 0.74 (SE = 0.08, *P =* 6.81 × 10’^-4^) between EAs and HAs. The cross-ancestry heritability estimates between AAs and HAs were too low to calculate ρ_pe_ between those ancestries.

In the supplementary analysis that excluded participants with a lifetime OUD diagnosis, we identified 3,400 SNPs in 101 LD-independent risk loci (Supplementary Table 6). Of these, 87 were GWS, 13 were *p* < 10’^6^ in the primary GWAS, and 18 were ancestry specific (17 in EAs and 1 in AAs) (Supplementary Tables 7 & 8).

### Single-nucleotide polymorphism heritability and enrichment

The proportion of variation in pain intensity explained by common genetic variants (h^2^_SNP_) was similar both for the full samples (AAs: 0.06 ± 0.009 and EAs: 0.08 ± 0.003) and the supplementary samples without OUD (AAs: 0.07 ± 0.009 and EAs: 0.08 ± 0.003) (Supplementary Table 9).

Partitioning the SNP heritability for pain intensity revealed significant tissue-group enrichment in central nervous system (CNS) (*P*⍰=⍰1.47⍰×⍰10^−12^), adrenal (*P*⍰=⍰8.97⍰×⍰10^−5^), liver (*P*⍰=⍰3.15⍰×⍰10^−4^), skeletal (*P*⍰=⍰8.50⍰×⍰10^−4^) and cardiovascular (P5H3.001) tissues (Figure 2A & B, Supplementary Table 10). In gene expression datasets derived from multiple tissues, we observed predominant h^2^_SNP_ effects in brain (*P*⍰=⍰2.87⍰×⍰10^−5^), including hippocampus (*P*⍰=⍰1.00⍰×⍰10^−4^) and limbic system (*P* = 1.15⍰×⍰10^−4^) (Figures 2C & D, Supplementary Table 11). SNP-based heritability in histone modification data also showed robust enhancer (H3K27ac and H3K4me1) and active promoter (H3K4me3 and H3K9ac) enrichments in brain tissues, including the dorsolateral prefrontal cortex (*P*⍰<⍰1.32⍰×⍰10^−4^), inferior temporal lobe *(P*⍰<⍰3.09⍰×⍰10^−4^), angular gyrus (*P*⍰=⍰8.42⍰×⍰10^−5^), and anterior caudate (*P*⍰=⍰1.12⍰×⍰10^−4^) (Figure 2E, Supplementary Table 12). Similar results were obtained for the partitioned heritability analysis of the supplementary GWAS (Supplementary Tables 11 & 12), though it also included significant expression effects in the cortex and cerebellum.

**Figure 2.**
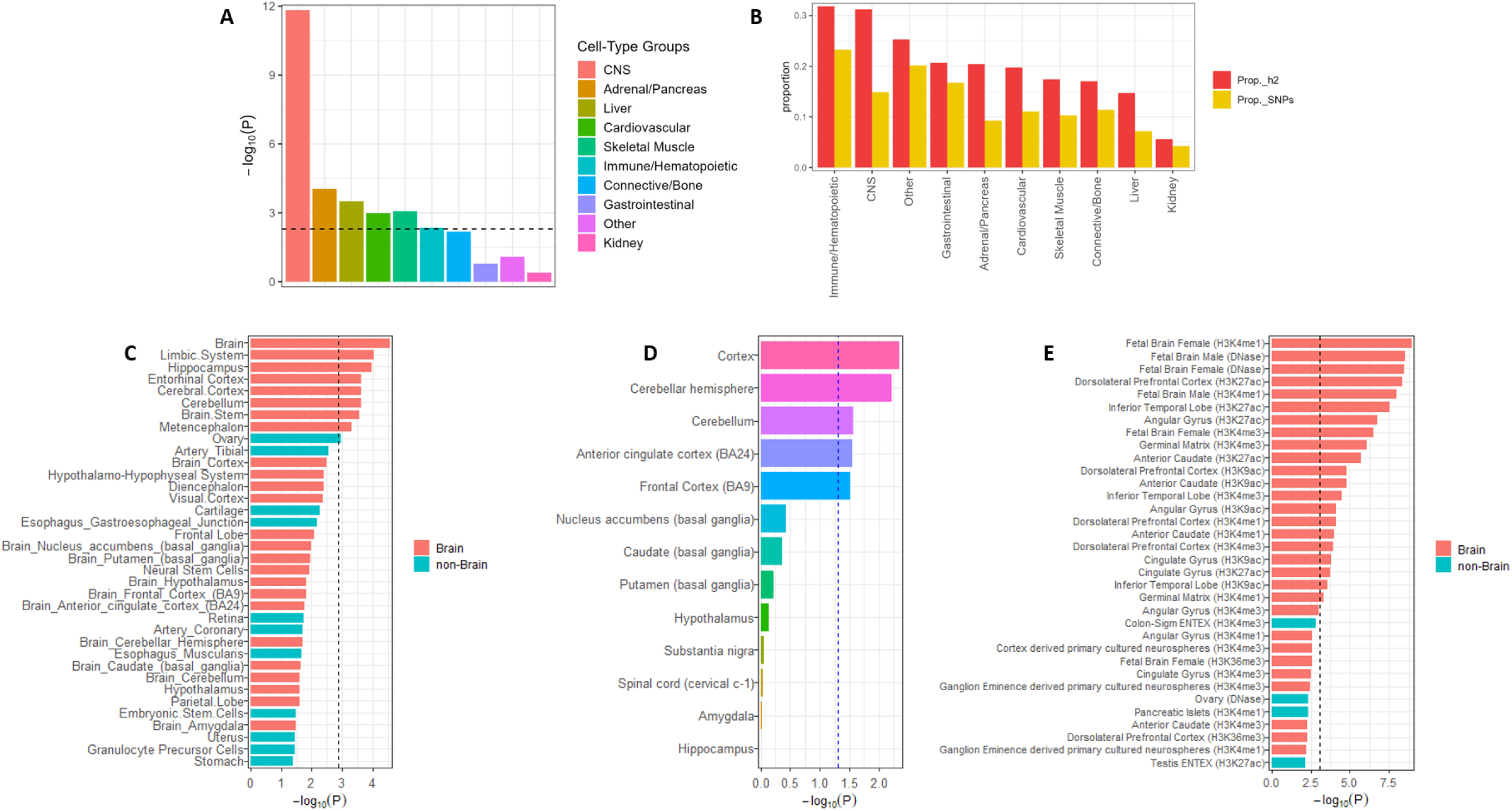
Enrichment of pain intensity in the brain. **A**, Partitioning héritability enrichment analyses using LDSC showing enrichment for pain intensity in the CNS, adrenal, liver, cardiovascular, and skeletal tissues. The dashed black lines indicate Bonferroni-corrected significance (*P*⍰<⍰O.OO5). **B**, Proportion of héritability shows robust enrichment for SNPs in brain and immune-related tissues. Héritability enrichment analyses for gene expression **(C & D)** and chromatin interaction (top 35 annotations are shown in E, see supplementary Table 12 for full details) using GTEx data show enrichment for pain intensity in brain regions previously associated with chronic pain. Bonferroni correction was applied within each tissue conditioned on the number of genes tested.

Although the SNP-based heritability and enrichment for the full and supplementary GWASs were similar, because the full sample yielded more risk loci, we based all downstream analyses (except genetic correlation [r_g_] analyses) on the GWAS results from that sample.

### Gene-set enrichment in tissue and cell types

To clarify the potential transcriptomic mechanism of each GWS pain locus, we mapped GWAS variants to genes via expression quantitative trait locus (eQTL) association in GTEx^61^ and assessed the tissue enrichment of mapped genes in FUMA^52^. After correcting for multiple testing (*P*⍰=⍰9.25⍰×⍰10^−4^) in the cross-ancestry and EA-specific GWASs, we uncovered significant transcriptomic enrichment only in brain tissues (Supplementary Figure 3). Consistent with previous findings of brain tissue enrichment across different pain phenotypes in EAs^22,25,27^, both our EA and cross-ancestry analyses showed notable enrichment in the cerebellum (cross-ancestry, *P*⍰=⍰2.48⍰×⍰10^−7^; EA, *P*⍰=⍰2.90⍰×⍰10^−6^), cerebellar hemisphere (cross-ancestry, *P*⍰=⍰4⍰×⍰10^−7^; EA, *P*⍰=⍰6.23⍰×⍰10^−6^), cortex (cross-ancestry, *P*⍰=⍰2.79⍰×⍰10^−6^; EA, *P*⍰=⍰3⍰×⍰10^−4^), and frontal cortex (cross-ancestry, *P*⍰=⍰2.82⍰×⍰10^−6^; EA, *P*⍰=⍰4.17⍰×⍰10^−4^) (Supplementary Figure 3). Among AAs there were no significantly enriched tissues (Supplementary Table 13).

To investigate enrichment at the level of the cell type in the EA GWAS results, we conducted FUMA cell-type specific analysis^67^ in a collection of cell types in 13 human brain sc-RNAseq datasets. After adjusting for possible confounding due to correlated expression within datasets using a stepwise conditional analysis, we detected jointly significant cell-type enrichments (proportional significance, PS > 0.5) for GABAergic neurons largely in the human adult mid-brain (*P*⍰=⍰l0.003 *β* = 0.206, s.e. = 0.075, PS 0.56) and to a lesser extent in the prefrontal cortex (*P*⍰=⍰O.O44, *β =* 0.045, s.e. = 0.016, PS 0.39) (Supplementary Table 14).

### Prioritization of candidate genes

To facilitate the biological interpretation and identification of druggable targets, we used a combination of MAGMA and fine-mapping, transcriptomic, proteomic, and chromatin interaction models to prioritize high-confidence variants and genes that most likely drive GWAS associations. Assigning SNPs to genes using physical proximity, MAGMA gene-based analyses^63^ identified 6 GWS genes in AAs, 203 in EAs, and 125 in the cross-ancestry results (Supplementary Figure 4, Supplementary Table 15), but none in HAs. MAGMA gene-set analysis^63^ using cross-ancestry GWAS results identified significantly enriched biological processes in catecholamine uptake (G0:0051944; Bonferroni *P*⍰=⍰0.019) and startle response (GO:0001964; Bonferroni *P*⍰=⍰O.O24). Negative regulation of synaptic transmission (GO:0050805; Bonferroni *P*⍰=⍰O.O16) was related to pain intensity in EAs (Supplementary Table 16).

For consistency with available reference data, we based the fine mapping procedure on EA GWAS results using 78 genomic regions (spanning 103 index variants) (Supplementary Table 17) defined by the maximum physical distance between the LD block of independent lead SNPs (Methods). Functional genomic prediction models used the full EA GWAS results (Supplementary Figure 1).

We fine-mapped the 78 regions using the Bayesian method implemented in FINEMAP^54^ (Methods). For each region with independent causal signals (Supplementary Table 17), credible sets of variants (PP > 0.5) were constructed to capture 95% of the regional posterior probability (k ≤ 5, Supplementary Table 18). Of these regions, 4 harbored 1 SNP (potentially indicating the causal variant), 20 regions 2 SNPs and 44 regions 3 or more SNPs (Supplementary Table 18). In total, FINEMAP prioritized 76 unique credible variants (N = 108, Figure 3A), including 26 independent lead SNPs and 18 novel pain loci (Figure 3B). Most (50/76) of the credible variants map to protein-coding genes and are mostly eQTLs (Supplementary Table 18), and five harbor missense variants, of which three *(ANAPC4, APOE*, and *SLC39A8)* are known pain loci^25,31^ and two *(RYR2* and *AKAP10)* are novel (Figure 3B). This small proportion of missense variants and high eQTL enrichment are consistent with an increased probability that the credible variants influence liability to pain intensity through gene expression modulation.

**Figure 3.**
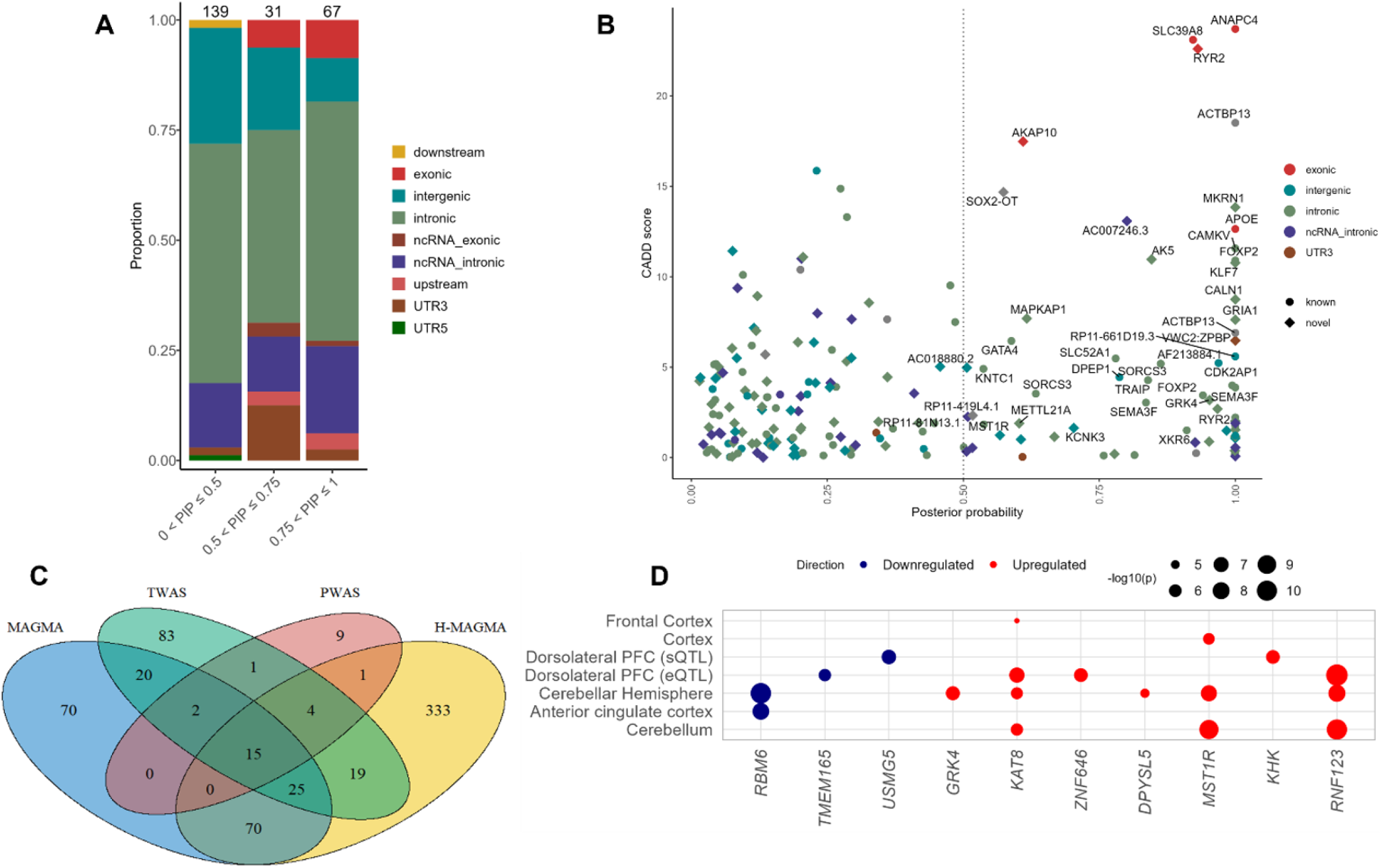
Gene prioritization for pain intensity. **A**, Genomic annotation of credible sets using FINEMAP shows enrichment largely in non-coding regions and to a lesser extent in exons. B, Annotation of known and novel credible genes. Dashed lines indicate posterior probability > 0.5. C, Number of overlapping genes across functional prediction models. D, Tissue enrichment of prioritized genes using SMR and GTEx data show enrichment in brain regions. Size of circle reflects -log_10_*P*. Bonferroni correction was applied within each tissue conditioned on the number of genes tested.

We performed TWAS and PWAS analyses to determine whether risk variants exert their effects via gene and/or protein expression. After correction for multiple testing, 196 unique genes (TWAS eQTL – 294, TWAS sQTL – 67 and PWAS – 32) were associated with pain intensity (Supplementary Tables 19 & 20). Of these, 69 represent novel associations (based on a window from the index GWAS locus > 1 MB). PWAS showed significant associations in the dorsolateral prefrontal cortex (dlPFC) that overlapped for 22 unique genes across multiple brain tissues in the TWAS (eQTL – 16, sQTL –8) (Figure 3C).

Chromatin interaction mapping using Hi-C data in adult and fetal brain identified 512 unique significantly interacting genes (*P*⍰=⍰2.84 *P*⍰×⍰10^−8^) (Supplementary Table 21), of which 60 are associated with all six chromatin annotations (Supplementary Figure 5) and 20 overlap with TWAS and/or PWAS findings, including *DPYSL5, KHK, MAPRE3, MST1R, NEK4, GNL3, GRK4, UHRF1BP1* and *VKORC1* (Figure 3C, Supplementary Tables 19, 20 & 21).

Based on concordant evidence from colocalization analyses in TWAS and PWAS (COLOC PP4 > 0.80), 104 unique genes (TWAS eQTL – 139, TWAS sQTL – 20 and PWAS – 14) were putatively causal for pain intensity (Supplementary Tables 19 & 20), of which 10 (including *DPYSL5, GRK4, KHK* and *MST1R)* were validated by SMR analysis (PHEIDI > 0.05) (Figure 3D, Supplementary Table 22). Among the 104 genes, 6 *(CHMP1A, GRIA1, GRK4, MST1R, STMN3* and *TRAF3)* captured 50% or more of the FINEMAP posterior probability (Supplementary Table 18). Notably, the *MST1R* intronic locus (rs9815930), which is in a credible set that harbors four other variants in high LD with the novel index variant rs2247036 (nearest gene – *TRAIP)* (Supplementary Figure 6), displayed the most robust causal effects from COLOC and SMR in more than one brain tissue (Figure 3D).

We also explored enrichment of causal genes and proteins in the dorsal root ganglia (DRG), which are important for transduction of nociceptive signals from the periphery to the CNS. None of the causal genes or proteins (N = 104) were enriched in human or mouse DRG (DRG enrichment score > 0.5) (Supplementary Figure 7A). Supporting results from TWAS and PWAS, 63 unique genes (human – 38 and mouse-49) were primarily enriched in the CNS, of which 22 (including *GRK4, GRIA1, MAPRE3, NEK4, STMN3* and *TRAF3)* showed common enrichment patterns across species (Supplementary Figure 7B).

Integrating FINEMAP, colocalization and SMR prioritized 156 high-confidence genes underlying the pain intensity GWAS association, of which 5 are exonic and missense (Supplementary Table 23), and 151 exert their effect via gene or protein expression.

### Phenotypic correlates of pain intensity

As expected, the strongest positive genetic correlations of pain intensity were with other pain phenotypes (e.g., multisite chronic pain *r*_g_=0.789, osteoarthritis *r*_*g*_*=0*.*710*, neck/shoulder pain *r*_g_=0.669, back pain *r*_g_=0.697, hip pain *r*_g_=0.729, knee pain *r*_g_=0.637; Figure 4A). Of 72 medical, anthropometric, or psychiatric traits associated epidemiologically with pain severity and mortality, 56 were significantly genetically correlated with pain intensity in EAs (Bonferroni *P <* 5.62 *P*⍰×⍰10’^4^) (Figure 4A, Supplementary Table 24).

**Figure 4.**
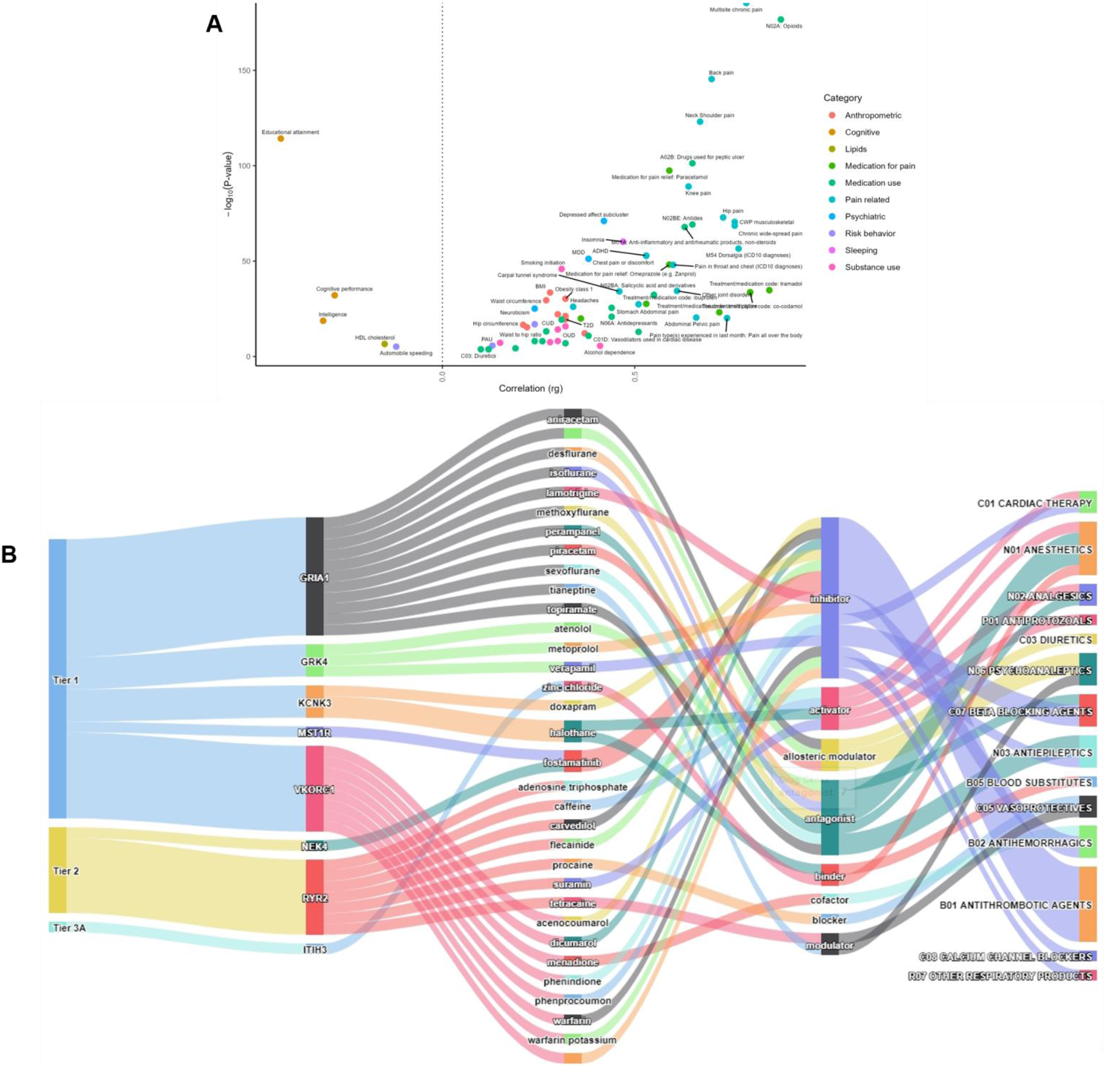
Genetic correlation and drug repurposing. **A**, Genetic correlation for pain intensity using LDSC. All points passing Bonferroni correction (Bonferroni correction threshold = 5.62⍰×⍰10’^4^ [0.05/89]) are plotted. The color of the circle indicates the phenotypic category. B, Druggable targets and drug interactions for 8 credible genes associated with pain intensity. For a full list of credible drug targets see Supplementary Table 30.

Notably, the liability to pain intensity was significantly positively genetically correlated with neuroticism, depression, insomnia, a variety of smoking-related measures, cannabis use disorder (CUD), alcohol dependence, OUD, and overweight and obesity (Figure 4A). As in prior studies^24^’^29^’^89^, pain intensity was significantly negatively correlated with educational attainment, cognitive performance, intelligence, and age of smoking initiation (Figure 4A). Relevant to drug repurposing, pain intensity was also positively correlated with the use of a variety of analgesic and anti-inflammatory drugs (Figure 4A). We also found significant *r*_g_s with pain intensity for several medical conditions and health outcomes in the UKBB (including genitourinary disease, chronic bronchitis, angina, etc., Bonferroni *P <* 3.72 *P*⍰×⍰10^−5^, Supplementary Table 25). In AAs, pain intensity was positively genetically correlated with PTSD-related features (e.g., re-experiencing, hyperarousal) and nominally associated *(p<* 0.05) with substance use traits (e.g., maximum alcohol intake and smoking trajectory, Supplementary Table 26).

In the Yale-Penn sample, we calculated PRS for 4,922 AAs and 5,709 EAs. Among AAs, none of the associations survived Bonferroni correction, likely due to the smaller discovery sample than for EAs (Supplementary Figure 8, Supplementary Table 27). In EAs, PheWAS identified 147 phenotypes, including 107 in the substance-related domain (40 opioid-related, 30 cocaine-related, 20 tobacco-related, 12 alcohol-related, and 6 cannabis-related) and 39 in other domains (9 medical, 18 psychiatric [9 PTSD, 5 ADHD, 2 conduct disorder, and 2 antisocial], 7 early childhood environmental, and 5 demographic phenotypes) that were significantly associated with the pain PRS (Supplementary Figure 9, Supplementary Table 27). The most significant findings were a negative association of the pain severity PRS with educational attainment (*P*⍰=⍰2.39⍰×⍰10^−26^) and a positive association with the Fagerström Test for Nicotine Dependence (*P*⍰=⍰4.71 ×⍰10^−256^). Opioid dependence was also positively associated with the pain PRS (*P*⍰=⍰3.87 ×⍰10^−12^), and remained significant when using a PRS based on the supplementary GWAS that excluded individuals with an OUD diagnosis (OR - 1.27, *P*⍰=⍰1.35 ×⍰10^−6^).

In PMBB, we calculated PRS for 10,383 AAs and 29,355 EAs. In AAs, no association with the pain PRS survived Bonferroni correction (Supplementary Figure 10, Supplementary Table 28). In EAs, the pain severity PRS was associated with 63 phenotypes, including 7 pain phenotypes and 6 psychiatric disorders (i.e., substance-, depression-, and anxiety-related traits). Other phenotypic categories with associations with the pain severity PRS were circulatory system (n = 1), infectious diseases (n=4), endocrine/metabolic (n=8), genitourinary (n=2), musculoskeletal (n=3), and neoplasms (n=4). The most significant findings were positive correlations with obesity (*P*⍰=⍰1.97⍰×⍰10^−45^) and tobacco use disorder (*P*⍰=⍰1.55⍰×⍰10^−24^) and a negative association with benign neoplasm of skin (*P*⍰=⍰2.67⍰×⍰10^−26^) (Supplementary Figure 11, Supplementary Table 28).

Two-sample MR between genetically correlated traits (N = 16) and pain intensity yielded 10 traits with evidence of causal association, 8 of which were bidirectional (Supplementary Table 29). Genetically predicted higher opioid use (N02A), depressed affect subcluster, major depressive disorder, neuroticism, use of drugs to treat peptic ulcer, and smoking cessation (coded as current smoking) had a significant positive bidirectional causal effect with pain intensity, whereas educational attainment and cognitive performance had a significant negative bidirectional causal effect (Supplementary Table 29). Further, increased risk of pain intensity positively predicted smoking initiation and cigarettes per day.

### Genetically inferred drug repurposing

Of the 156 genes in EAs with evidence supporting causality from fine-mapping and functional genomic prediction, 20 were present in the druggable genome database^76^ (Supplementary Table 30). Of these druggable candidate genes, 11 (including *GRIA1, GRK4* and *MST1R)* are tier-1 candidates, which includes targets of licensed drugs and drugs in clinical trial, 4 genes (e.g., *NEK4* and *RYR2)* are in tier 2, and 4 are in tier 3 (Supplementary Table 30). Within tier 1, drugs that interact with *GRK4* (a credible pain gene locus in moderate LD with the novel index variant *NOP14*\*rs71597204 – Supplementary Figure 12) are beta-blockers (atenolol and metoprolol) and a calcium-channel blocking agent (verapamil) (Figure 4B), which have analgesic effects in osteoarthritis^90,91^ and migraine^92^. Another tier-1 candidate gene – *GRIA1* – is targeted by anesthetics (sevoflurane, isoflurane, desflurane), antiepileptics (topiramate, perampanel), analgesics (methoxyflurane), psychoanaleptics (piracetam, aniracetam), and a diuretic (cyclothiazide) (Figure 4B). Drug classes for pain intensity also included anti-hemorrhagic agents (e.g., fostamatinib [tier 1: *MST1R* and *FYN;* tier 2: *NEK4]* and menadione *[VKORC1])* (Figure 4B, Supplementary Table 30).

Of the 7 genes associated with pain intensity in AAs, *PPARD*, which harbors the new genetic signal discovered in this study, is a tier-1 druggable candidate with 30 interacting drug classes (Supplementary Table 30). The *PPARD* negative modulator sulindac is an approved non-steroidal antiinflammatory and antirheumatic drug used to treat osteoarthritis.

### Discussion

*We* conducted the largest multi-ancestry, single-sample GWAS of pain intensity to date, comprising 112,968 AA, 436,683 EA, and 48,688 HA individuals. Cross-ancestry analyses identified 125 independent risk loci, of which 82 have not previously been associated with any pain phenotype. Although prior GWASs for chronic pain phenotypes have identified 99 loci^23-27^’^32^, the study samples have largely been limited to EA individuals. The diversity of the MVP sample enabled us to identify novel association signals in both AAs (*PPARD*\*rs9470000) and HAs (nearest genes *RAU6*-*461P*\*rsl46862033, *RNU6-741P*\* rsl019597899).

Findings from gene set analysis, tissue enrichment, and cell-type specificity highlight novel biological pathways linking genetic variation to the etiopathology of pain. These functional analyses all implicate the brain, providing genetic support to the current understanding of the pathophysiology of pain severity^93^. Genes predominantly expressed in the CNS, particularly in the cerebellum, cerebellar hemisphere, and cortex region, rather than in the DRG, appear to play a salient role in modulating the intensity of pain, consistent with prior associations of sustained chronic pain intensity with increased activity in these brain regions^94-96^. Our findings are also consistent with prior reports^24,25,97,98^ of enriched gene expression in brain that contribute to pain intensity in a dose- and time-dependent manner and may involve specific neuronal processes in brain regions implicated in emotional processing^93^. Evidence that GABAergic neurons are cells of specific interest is a key novel finding. GABA has long been implicated in the modulation and perception of pain^99-101^ and previous work has implicated specific GABAergic activity in the midbrain as a modulator of pain and anxiety^102^. Altered GABA levels have been reported in individuals with various types of pain^103,104^, and have been associated with greater selfreported pain^105^. Targeting GABA functioning (e.g.,^106^), particularly in the brain regions enriched for pain intensity, may represent a novel therapeutic strategy.

Eleven of 156 prioritized genes encode druggable small molecules that are targets of licensed drugs or those in clinical trials, representing drug repurposing opportunities for treating chronic pain. We highlight *GRK4* and *GRIA1*, each with at least three lines of evidence supporting their involvement in chronic pain. *GRK4* encodes G protein-coupled receptor kinase 4 and has been linked with hypertension^107^, which is associated with chronic pain at the population level^108,109^. Of note, *GRK4* showed significant upregulation in the cerebellar hemisphere, fine maps to an intronic variant with > 95% PP, and is a target of beta-blockers. The use of beta-blockers has been associated with reduced osteoarthritis pain scores, prescription analgesic use^90^ and consultations for knee osteoarthritis, knee pain, and hip pain^91^. *GRIA1* encodes an ionotropic glutamate receptor subunit, an excitatory neurotransmitter receptor at many synapses in the CNS. Loss-of-function mutations in *GRIA1* are linked to neurodevelopmental impairments^110,111^. The *GRIA1* antagonist sevoflurane reduced pain in patients suffering from chronic venous ulcer^112^. However, clinical trials of topiramate (another drug target for *GRIA1)* for treating neuropathic chronic pain are inconclusive^113^. Research on the mechanisms that underlie the biology of these potential drug targets for *GRK4* and *GRIA1* and their effects on the onset and severity of chronic pain are warranted.

Pain intensity was strongly genetically correlated with other chronic pain phenotypes. Corroborating existing epidemiological studies on the comorbid nature of different pain conditions^33^, the strongest genetic correlations of pain intensity were with multisite chronic pain, followed by pain in specific bodily locations. In line with previous observations in GWASs of other pain-related phenotypes^24^’^25^’^27^’^28^’^89^, there were also positive genetic correlations of pain intensity with psychiatric disorders, substance use and use disorders, and anthropometric traits.

PheWAS findings in both the Yale-Penn sample - enriched for individuals with substance-related traits - and the PMBB - comprising a medical population - were prominent in EAs. These findings underscore the important influence of co-occurring substance-related, psychiatric, and medical pathology and educational achievement on the intensity of the pain experience. In contrast, the PRS generated from the pain intensity discovery sample in AAs yielded few associations in either of the target samples, which underscores the need for larger non-European samples to elucidate the genetic architecture of pain intensity.

Two-sample MR analysis supported causal associations between pain and multiple traits.

Smoking has previously been associated with greater pain intensity, but studies can be confounded by socioeconomic factors, and a bi-directional relationship has been proposed^114^. Here, we show evidence for a causal relationship of pain on the number of cigarettes smoked per day, smoking initiation, and smoking cessation. In line with previous findings^25^’^33^’^35^’^36^, pain intensity had a bidirectional causal effect on the risk of both depression and neuroticism, suggesting that greater pain could predispose individuals to increased risk for these psychiatric disorders and vice versa. Supporting the positive genetic correlation between opioid use and pain intensity, MR showed evidence of a bidirectional causal effect between pain intensity and opioid use.

Our findings underscore the complex nature of pain intensity, with the hundreds of genetic loci contributing to the experience of pain identified here and in prior studies reflecting a substantial genetic contribution to pain-related traits. The evidence adduced here of pleiotropy of pain intensity with psychiatric traits such as neuroticism and depression reflects the contribution of non-physical factors to the experience of pain intensity. This is consistent with the observed significant tissue-group enrichment in CNS, the predominant gene expression findings in brain (including the hippocampus and limbic system), and the SNP-based enhancer enrichments in histone modification in brain tissues (including the dorsolateral prefrontal cortex, inferior temporal lobe, angular gyrus, and anterior caudate).

A limitation of the present study concerns the NRS phenotype. Although such a quantitative trait is more informative than a binary one (e.g., the presence of a specific pain diagnosis), it is based on subjective report. However, because the subjective experience of pain is a key defining feature of the clinical phenomenon^1,115^ the phenotype has high public health significance. Pain scores recorded by clerks and nurses in the clinical setting may consistently under report the patient’s response. In earlier work that compared self-reported pain from a direct patient survey to scores recorded in a VA clinical setting^117^, we found that, despite lower scores recorded in the clinic the two reports correlated well. Nonetheless, the imprecise measurement of pain intensity likely yields lower power for gene discovery. The routine assessment of pain severity provided a very large number of pain scores, which we reduced by taking the median of medians for each individual as a trait for GWAS. In subsequent analyses, we plan to evaluate alternative methods for characterizing pain severity (e.g., pain trajectories). Another limitation is that our sample comprises predominantly male veterans, which in view of well demonstrated sex differences in the experience and frequency of pain^26^, limits the application of the findings to the general population. Finally, although our sample was more diverse than prior GWAS of pain traits, analyses in the AA and HA samples were underpowered.

Despite these limitations, the large MVP sample and informative quantitative trait measured repeatedly within subjects enabled us to generate a proxy for chronic pain and identify many novel loci contributing to the trait. Downstream analyses localize the genetic effects largely to four CNS regions and using available single-cell RNAseq data specifically to GABAergic neurons. Combined with drug repurposing findings that implicate 20 druggable targets, the study provides a basis for studies of novel, non-opioid medications for use in alleviating chronic pain.

## Data Availability

All genotype data and summary phenotype data from the Million Veteran Program GWAS will be posted on dbGaP upon acceptance of the manuscript in a peer-review journal

## Data Availability

The full summary statistics from the meta-analyses will be available through dbGaP upon publication.

## Code Availability

Imputation was performed using Minimac3 (https://genome.sph.umich.edu/wiki/Minimac3). GWAS was performed using PLINK2 (https://www.cog-genomics.org/plink2). Meta-analyses were performed using METAL (https://genome.sph.umich.edu/wiki/METALDocumentation). GCTA-COJ0 (https://cnsgenomics.eom/software/gcta/#Overview) was used for identification of independent loci. FUMA (https://fuma.ctglab.nl/) was used for gene association, functional enrichment and gene-set enrichment analyses. Transcriptomic and proteomic analyses were performed using FUSION (https://github.com/gusevlab/fusiontwas). Chromatin accessibility analyses were performed using H-MAGMA (https://github.com/thewonlab/H-MAGMA). LDSC (https://github.com/bulik/ldsc) was used for heritability estimation, genetic correlation analysis (also using the CTG-VL; https://genoma.io) and heritability enrichment analyses. Trans-ancestry genetic correlation was estimated using Popcorn (https://github.com/brielin/Popcorn). PRS analyses were performed using PRS-CS (https://github.com/getianlO7/PRScs). PheWAS analyses were run using the PheWAS R package (https://github.com/PheWAS/PheWAS). The MendelianRandomization R package (https://cran.r-project.org/web/packages/MendelianRandomization/index.html) was used for MR analyses.

## Acknowledgements

This work was supported by grants from the US Department of Veterans Affairs Biomedical Laboratory Research and Development Service (no. I01 BX003341 (to A.C.J. and H.R.K.)) and the VISN 4 Mental Illness Research, Education and Clinical Center (to H.R.K.); and NIH grants K01 AA028292 (to R.L.K.); and P30 DA046345 (to H.R.K.). The funders had no role in study design, data collection and analysis, decision to publish, or preparation of the manuscript. The views expressed in this article are those of the authors and do not necessarily represent the position or policy of the Department of Veterans Affairs or the US Government.

We acknowledge the Penn Medicine BioBank (PMBB) for providing data to generate polygenic risk scores and conduct PheWAS analyses and thank the patients of Penn Medicine who consented to participate in this research program. We would also like to thank the Penn Medicine BioBank team and Regeneron Genetics Center for providing genetic variant data for analysis. The PMBB is approved under IRB protocol# 813913 and supported by Perelman School of Medicine at University of Pennsylvania, a gift from the Smilow family, and the National Center for Advancing Translational Sciences of the National Institutes of Health under CTSA award number UL1TR001878.

This manuscript has been co-authored by UT-Battelle, LLC under Contract No. DE-AC05-00OR22725 with the U.S. Department of Energy. The United States Government retains and the publisher, by accepting the article for publication, acknowledges that the United States Government retains a non-exclusive, paid-up, irrevocable, world-wide license to publish or reproduce the published form of this manuscript, or allow others to do so, for United States Government purposes. The Department of Energy will provide public access to these results of federally sponsored research in accordance with the DOE Public Access Plan (http://energy.gov/downloads/doe-public-access-plan).

## Contributions

S.T conducted the main analyses and drafted the manuscript. R.V.S conducted phenotype-related analyses. Z.J and H.X conducted downstream analyses. D.S annotated gene findings. M.P.V and K.S helped conduct analyses. R.V.S, Z.J, H.X, D.S, E.H, M.P.V, K.S, K.X, J.G, D.A.J, C.T.R, M.C, E.S, and S.G.W helped to write the manuscript. A.C.J obtained funding to support the project and helped to write the manuscript. R.L.K supervised the analyses and helped to write the manuscript. H.R.K conceived the project, obtained funding to support it, and helped to supervise the analyses and write the manuscript. All authors reviewed and approved the final version of the manuscript

## Ethics declarations

HRK is a member of advisory boards for Dicerna Pharmaceuticals, Sophrosyne Pharmaceuticals, Enthion Pharmaceuticals, and Clearmind Medicine; a consultant to Sobrera Pharmaceuticals; the recipient of research funding and medication supplies from Alkermes for an investigator-initiated study; and a member of the American Society of Clinical Psychopharmacology’s Alcohol Clinical Trials Initiative, which was supported in the last three years by Alkermes, Dicerna, Ethypharm, Lundbeck, Mitsubishi, and Otsuka. HRK and JG are named as inventors on PCT patent application #15/878,640 entitled: “Genotype-guided dosing of opioid agonists,” filed January 24, 2018. ES is a full-time employee of Regeneren Pharmaceuticals. The other authors have no disclosures to make.

