## Supplementary Figures 1-12 for "The genetic architecture of pain intensity in a sample of 598,339 U.S. veterans"

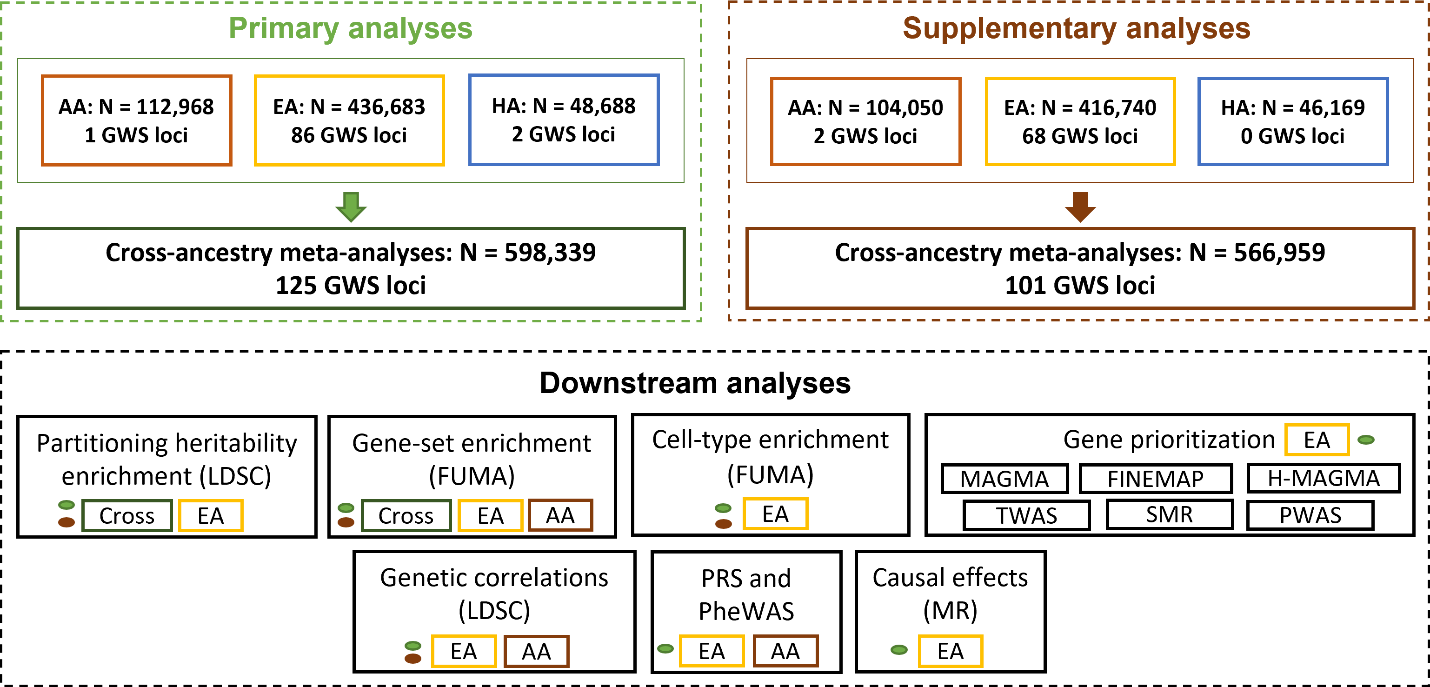

**Supplementary Figure 1. Overview of the study**. Top left: Primary GWAS analyses for pain intensity. Within ancestry GWAS for African American (AA), European American (EA) and Hispanic American (HA) followed by cross-ancestry meta-analysis. These results were used for all downstream analyses. Top right: Supplementary GWAS analyses for pain intensity. Bottom: Downstream analyses were conducted using the cross-ancestry, AA and EA GWAS results as indicated by color shadings: primary GWAS (green) and supplementary GWAS (brown)

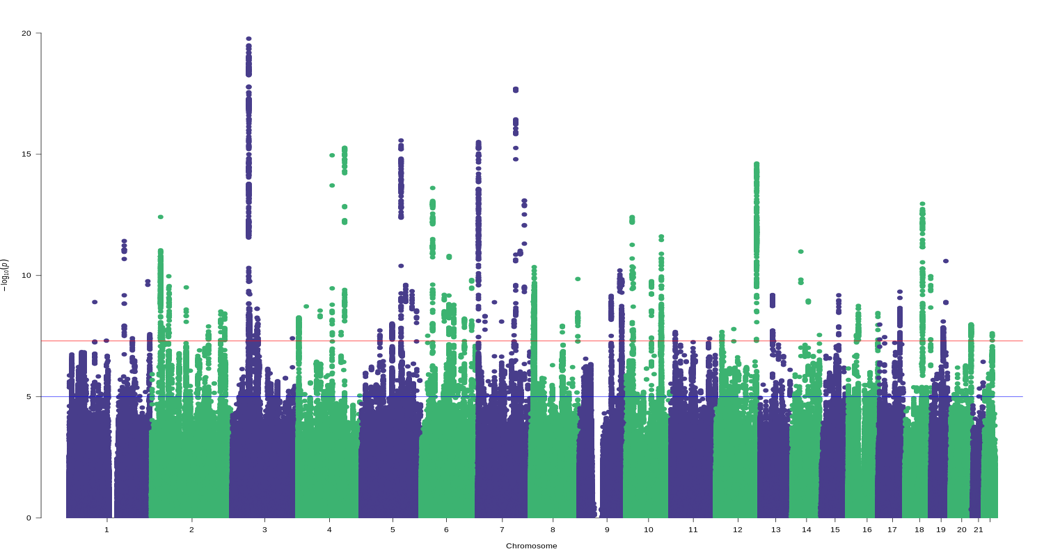

**Supplementary Figure 2. Manhattan plot for the pain intensity in European American GWAS analysis.** Identified 86 independent risk loci. The red line indicates GWS after correction for multiple testing (*P* < 5 × 10^−8^)

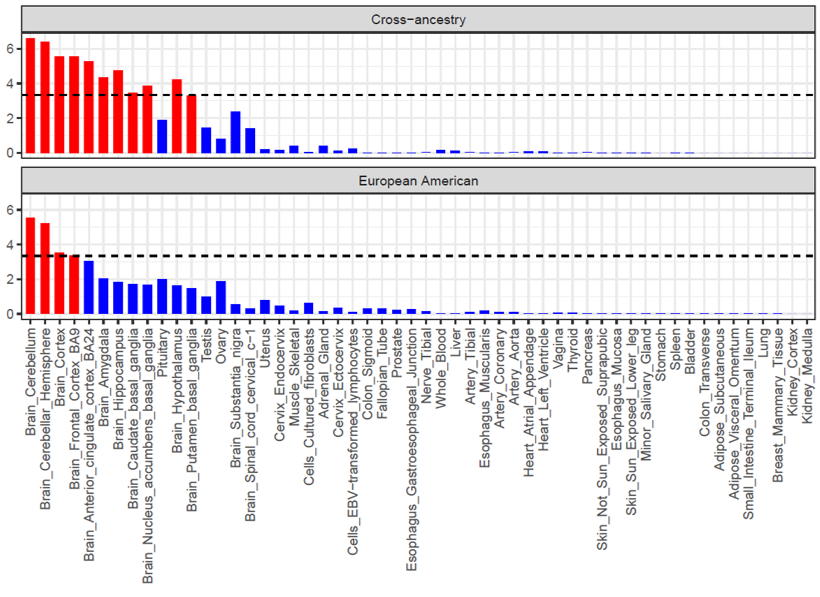

**Supplementary Figure 3. MAGMA tissue enrichment for the pain intensity in cross-ancestry and European American GWAS results.** Tissue enrichment analyses were conducted using FUMA. Bonferroni correction threshold (represented by the black dashed line) = 9.25 × 10^-4^ (0.05/54)

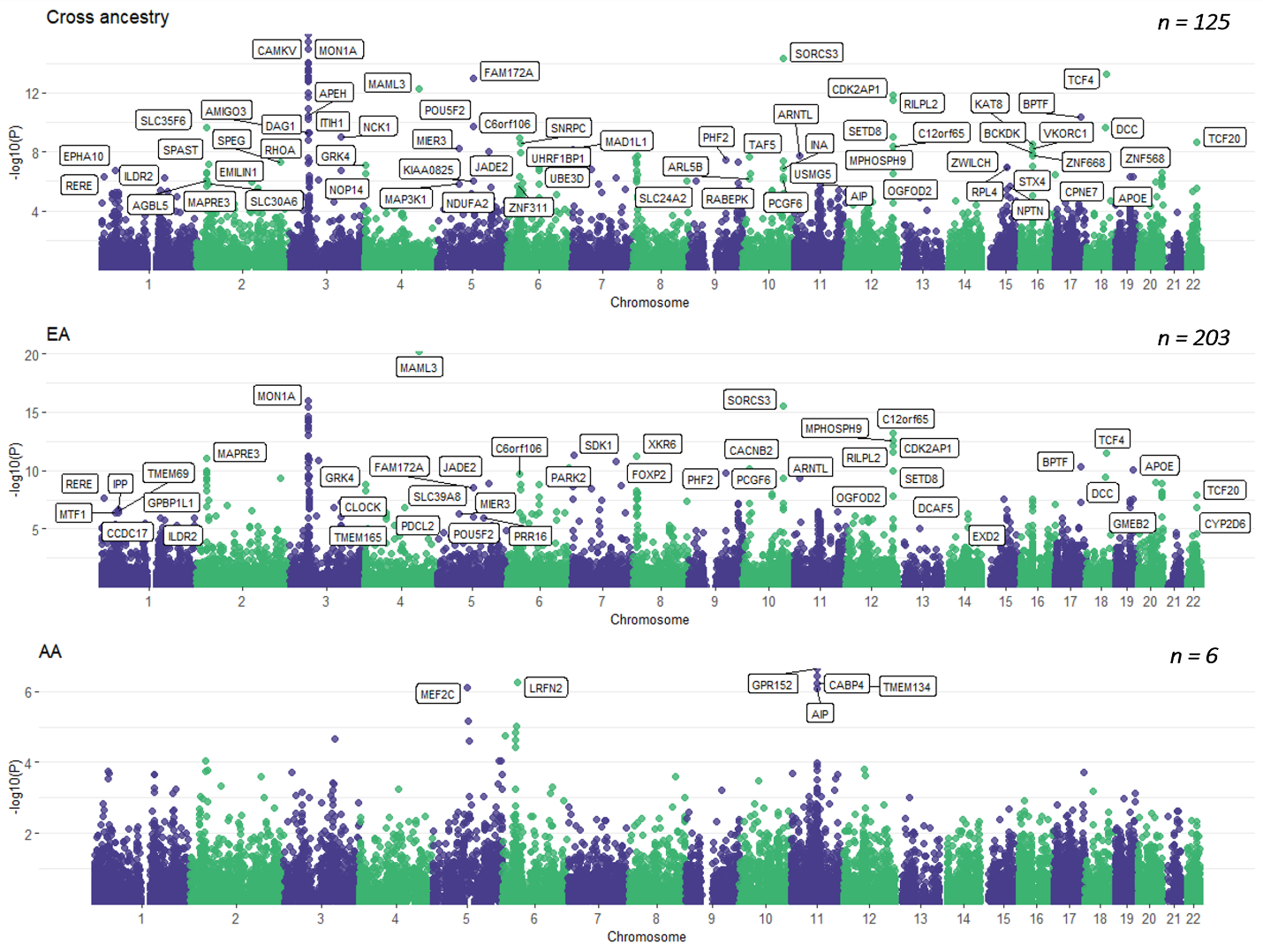

**Supplementary Figure 4. Gene-based Manhattan plots for cross-ancestry, European American and African American GWAS.** Gene-based association analyses were conducted using FUMA and genes that survive multiple correction are annotated. (Bonferroni *p* = 2.67 × 10^-6^ [0.05/18,702]).

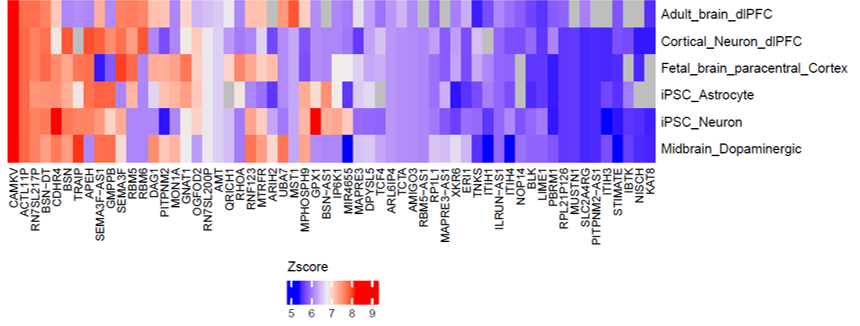

**Supplementary Figure 5. H-MAGMA Gene-Tissue pairs for pain intensity.** Genes that survive multiple correction across six tissues are annotated. (Bonferroni *p* = 2.84 × 10^-8^)

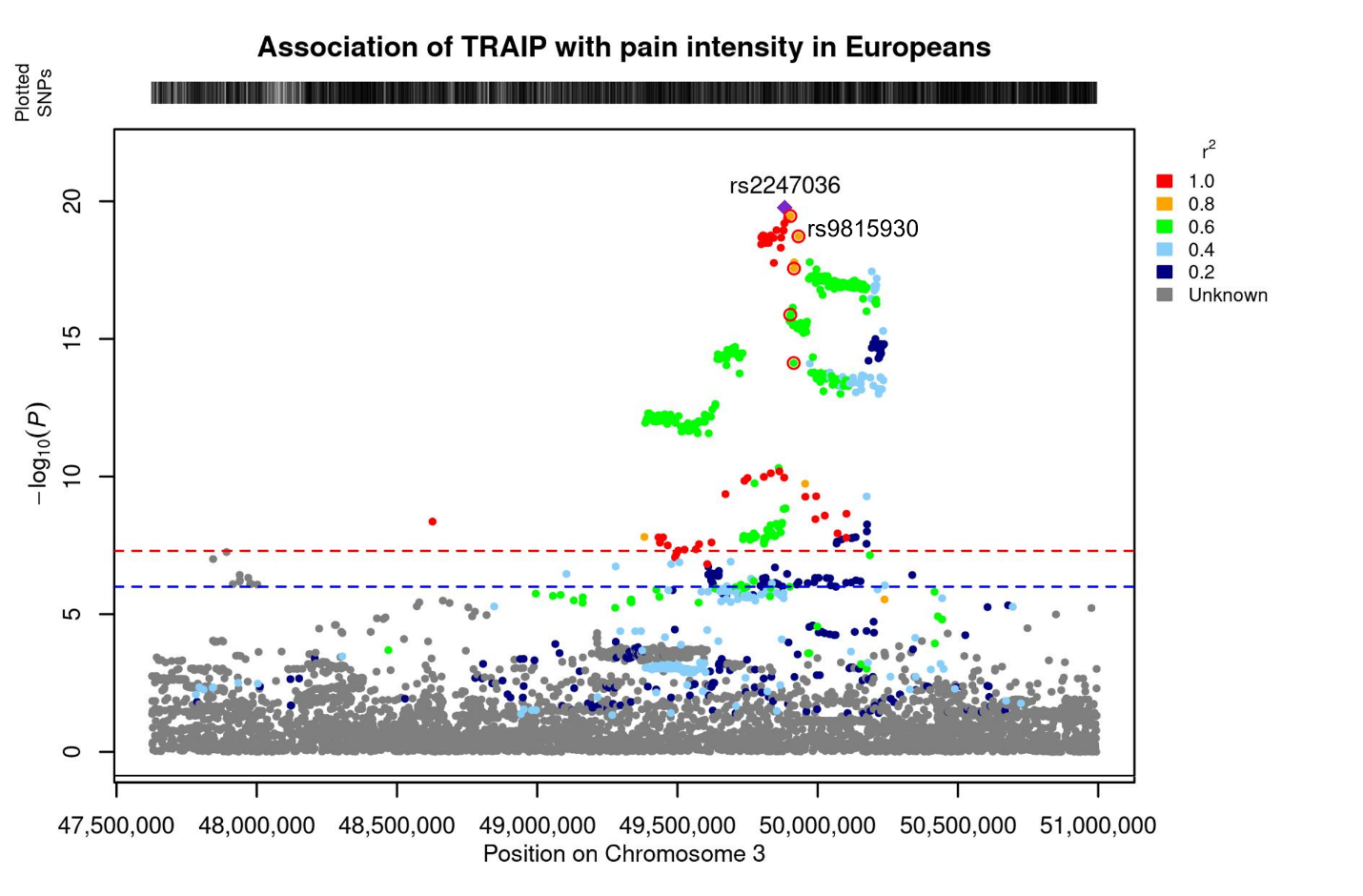

**Supplementary Figure 6. Regional plot for TRAIP*rs2247036 and MSTR1*rs9815930 on chromosome 3.** Credible locus prioritized by FINEMAP (PP > 0.5) are annotated with red rings. The *MSTR1**rs9815930 locus is in high LD (r2 > 0.8) with the lead variant *TRAIP**rs2247036.

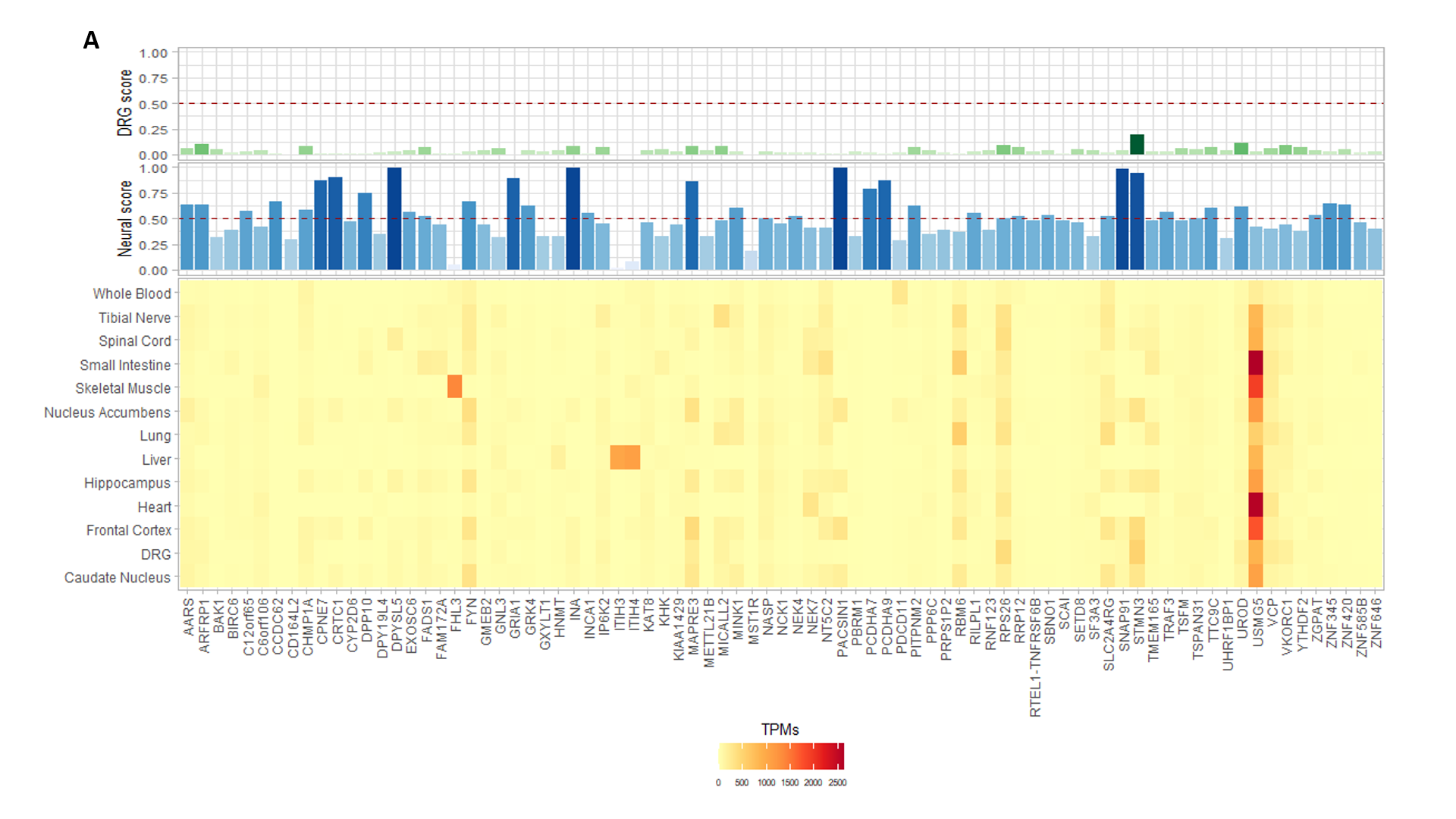

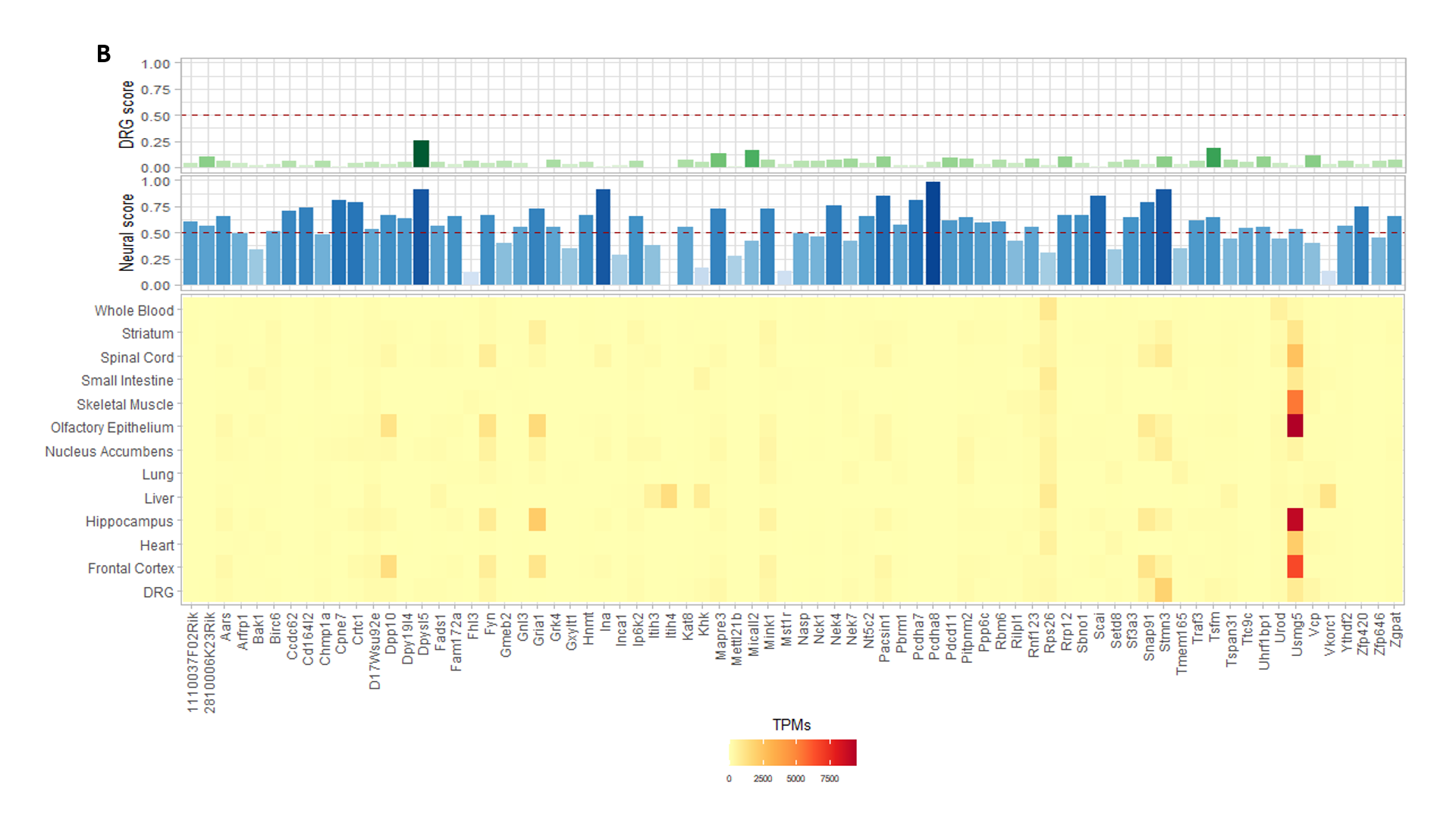

**Supplementary Figure 7. Identification of tissue enrichment gene expression patterns.** Causal genes and proteins that are differentially expressed in CNS and DRG in (A) humans and (B) mouse. DRG, dorsal root ganglia; TPM, transcripts per million.

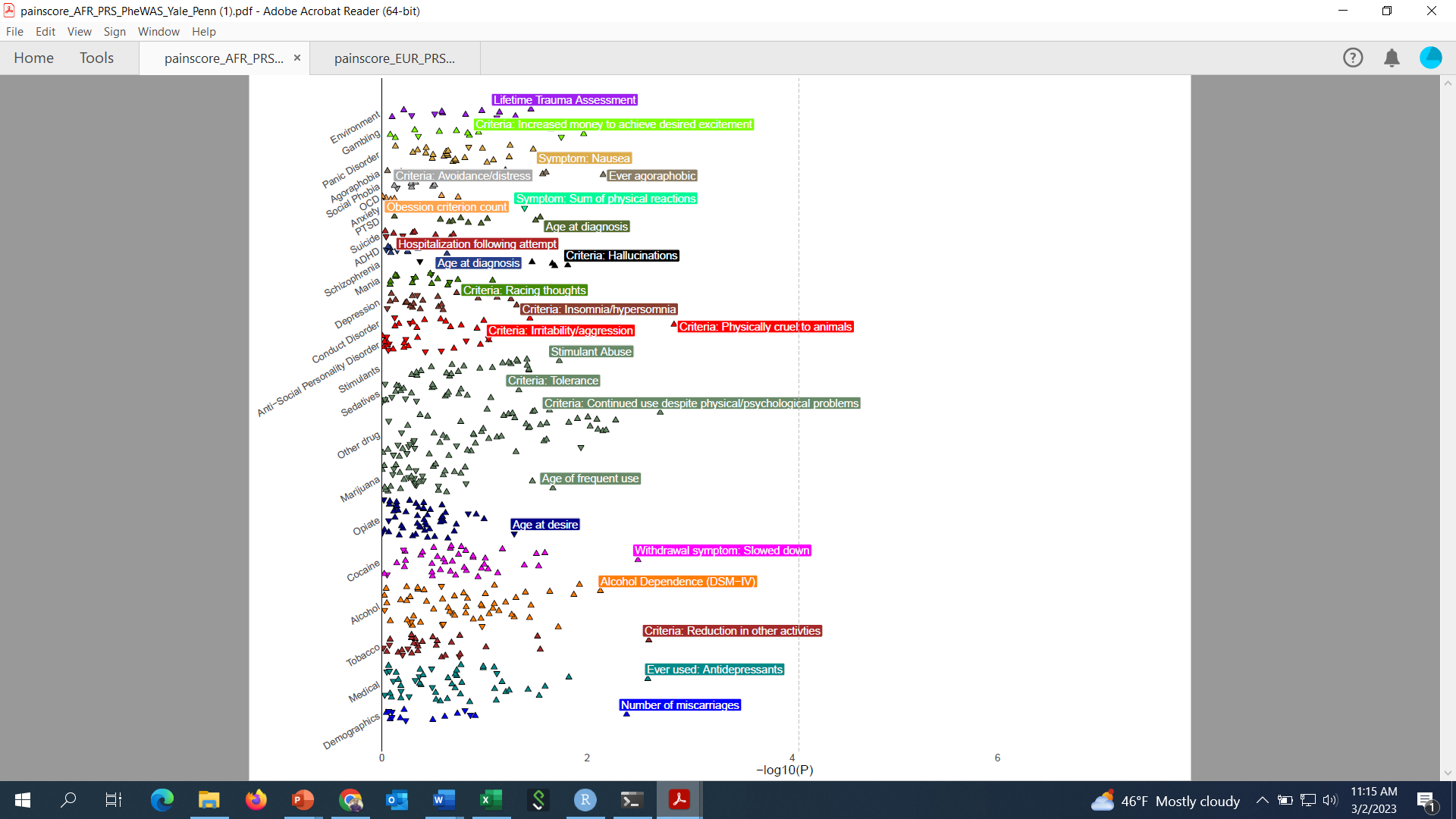

**Supplementary Figure 8. PheWAS in Yale-Penn AA individuals.** PheWAS plot for pain intensity PRS in EA individuals from Yale-Penn. No phenotypes passed Bonferroni correction (Bonferroni correction threshold = *P <* 7.83 × 10^-5^ (0.05/638).

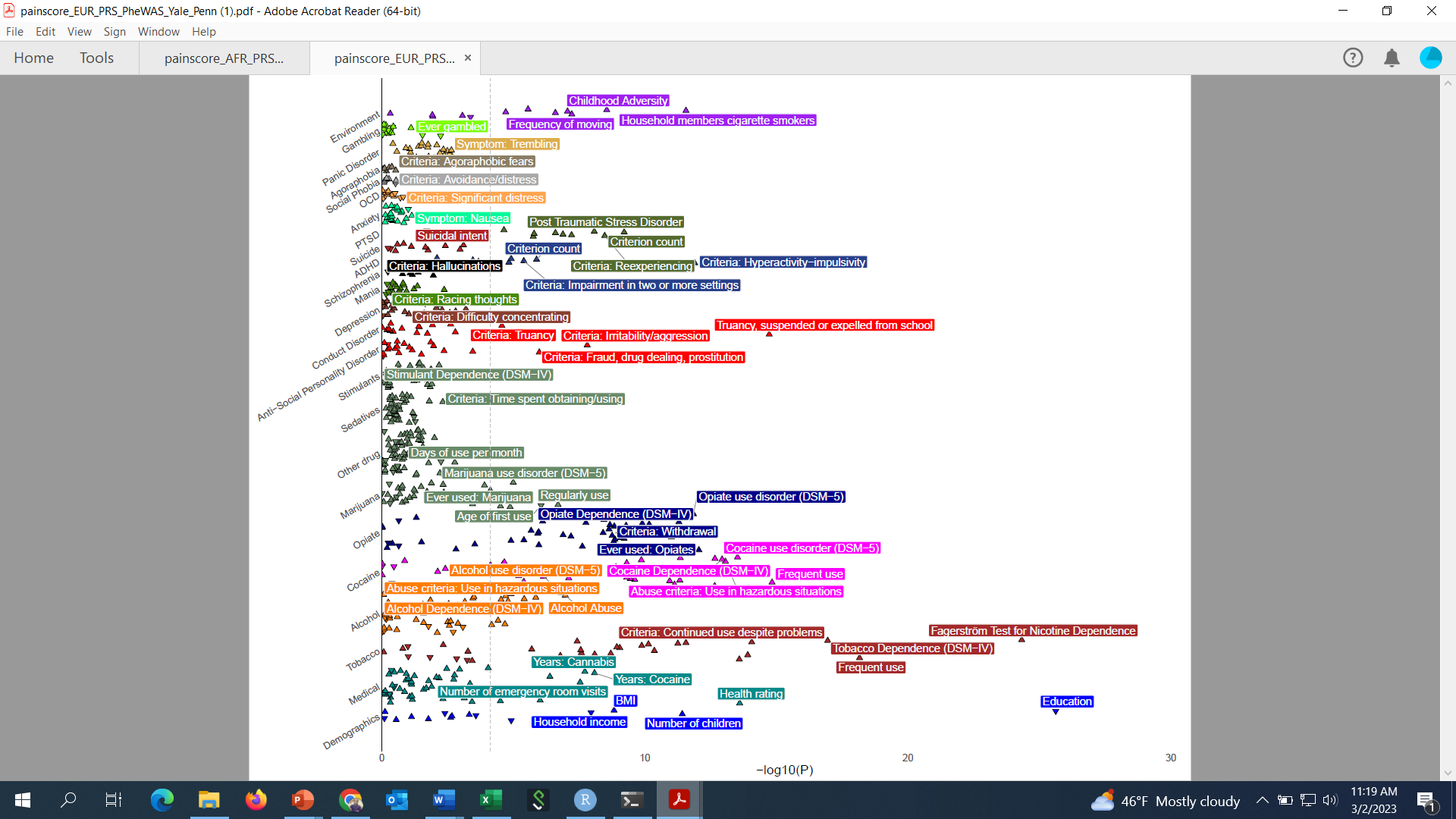

**Supplementary Figure 9. PheWAS in Yale-Penn EA individuals.** PheWAS plot for pain intensity PRS in EA individuals from Yale-Penn. Phenotypes that pass Bonferroni correction are annotated (Bonferroni correction threshold = *P <* 7.83 × 10^-5^ (0.05/638).

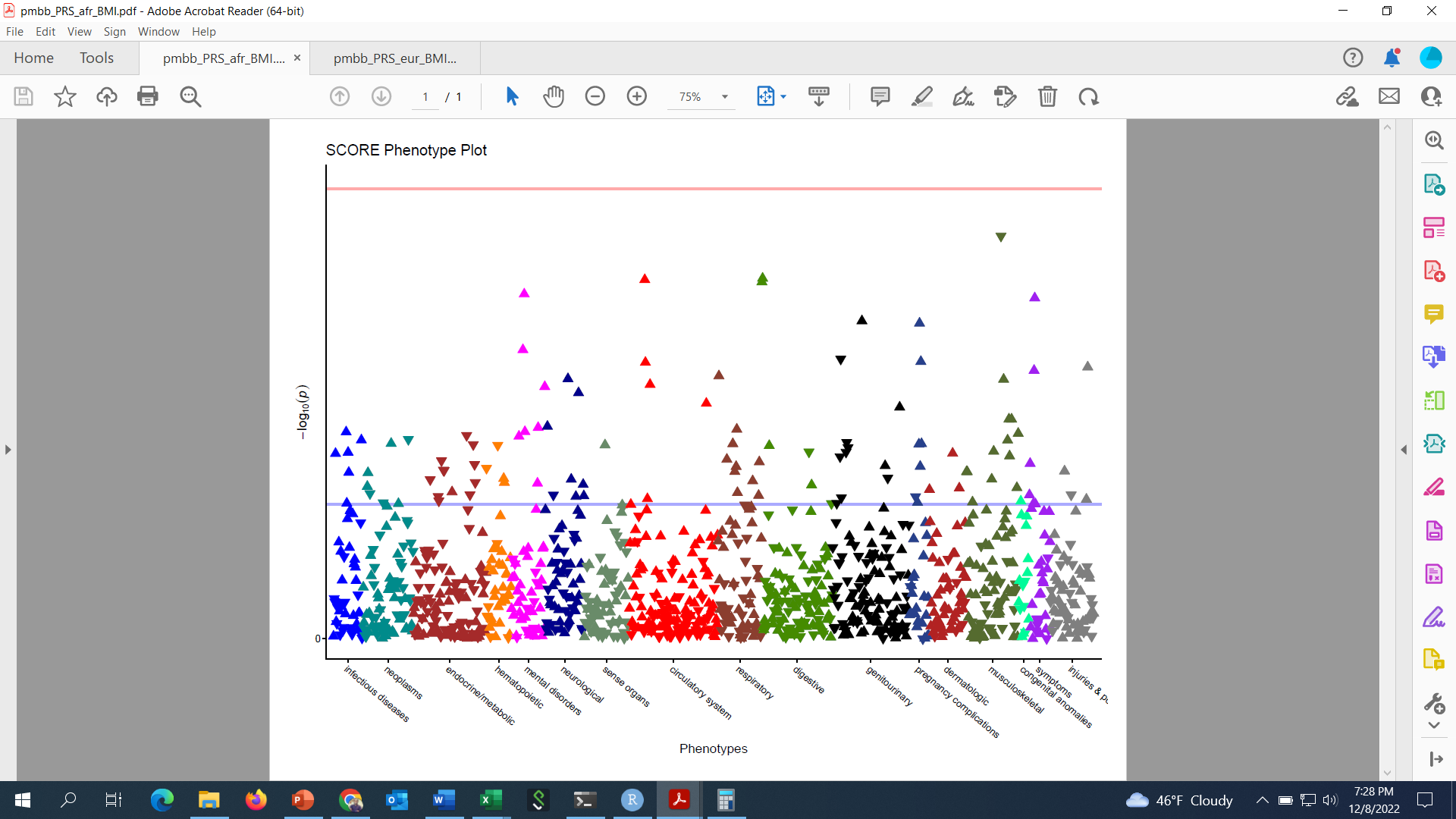

**Supplementary Figure 10. PheWAS in PMBB AA individuals.** PheWAS plot for pain intensity PRS in AA individuals from PMBB. No phenotypes pass Bonferroni correction are annotated (Bonferroni correction threshold = *P <* 3.68 × 10^-5^ (0.05/1360).

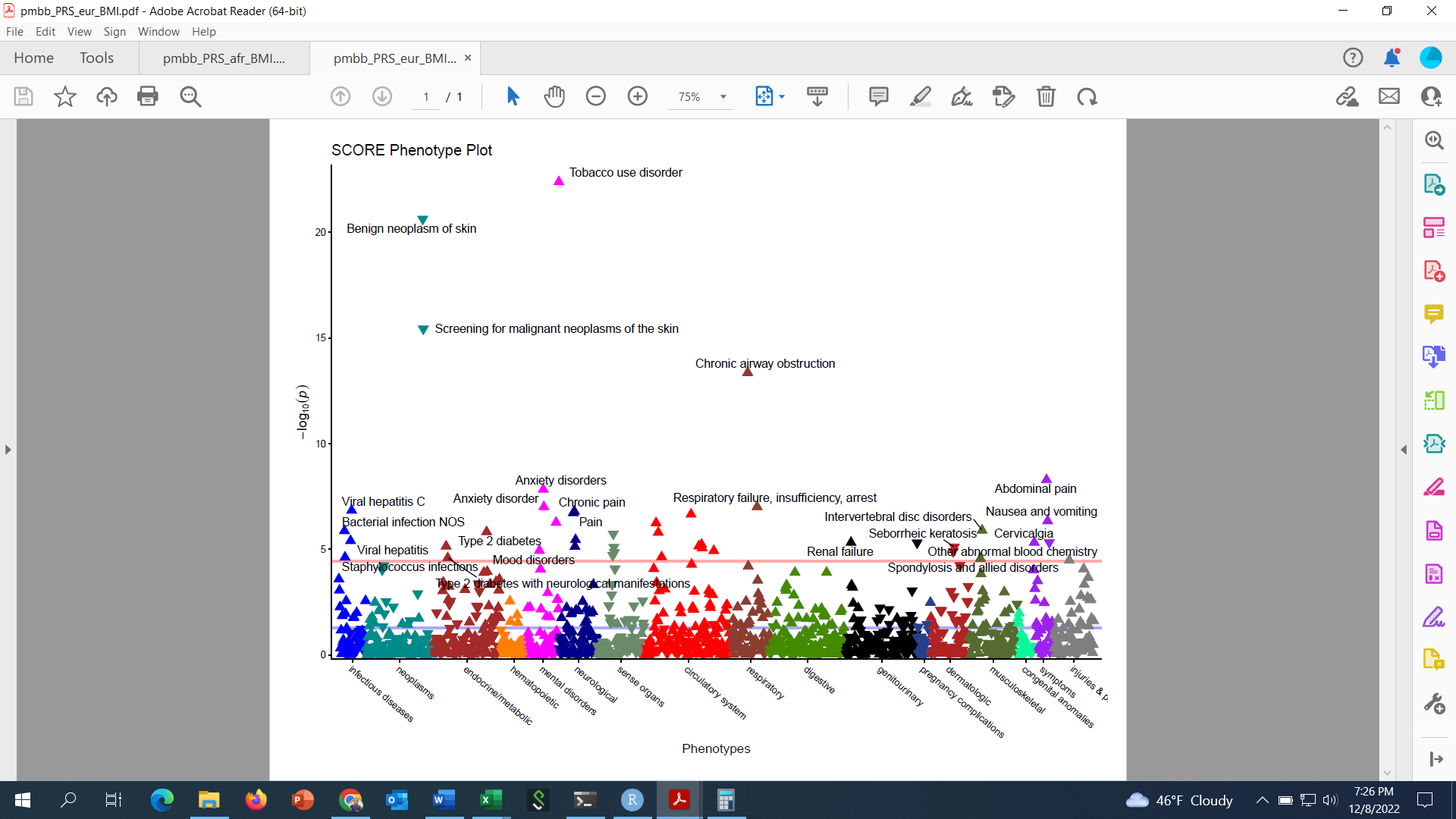

**Supplementary Figure 11. PheWAS in PMBB EA individuals.** PheWAS plot for pain intensity PRS in EA individuals from PMBB. Phenotypes that pass Bonferroni correction are annotated (Bonferroni correction threshold = *P <* 3.68 × 10^-5^ (0.05/1360).

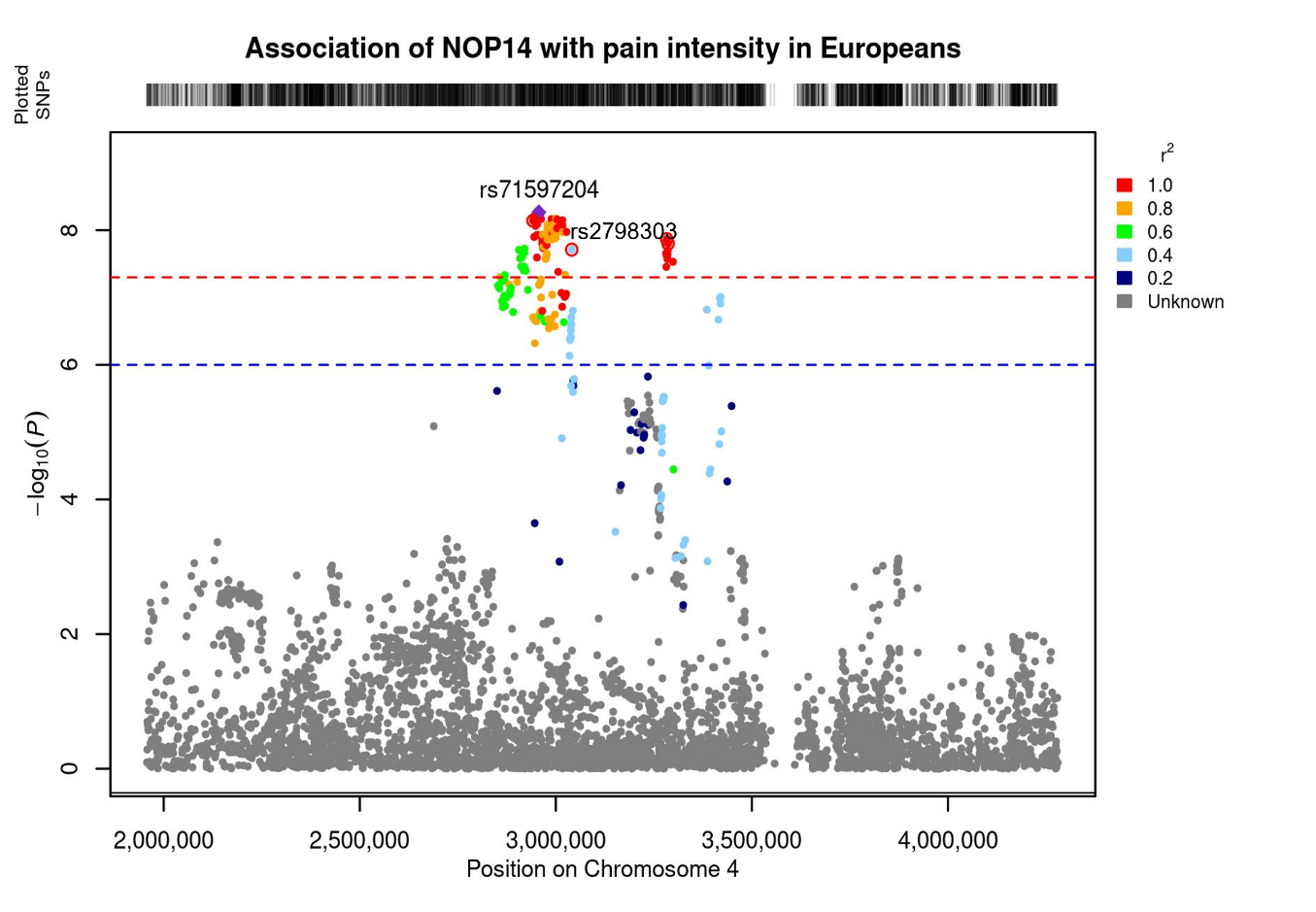

**Supplementary Figure 12. Regional plot for *NOP14**rs71597204** **and *GRK4**rs2798303 on chromosome 4.** Credible locus prioritized by FINEMAP (PP > 0.5) are annotated with red rings. The *GRK4**rs2798303 locus is in moderate LD (r2 > 0.4) with the lead variant *NOP14**rs71597204.
